# Expanded map of genomic imprinting reveals insight into human disease

**DOI:** 10.1101/2025.09.15.25335770

**Authors:** Craig Smail, Warren A. Cheung, Boryana Koseva, Adam F. Johnson, Chengpeng Bi, Carl F. Schreck, Michael Lydic, Kristin Holoch, Elena Repnikova, John Herriges, Courtney Marsh, Isabelle Thiffault, Tomi Pastinen, Elin Grundberg

## Abstract

Genomic imprinting involves parent-of-origin effect (POE) of regulatory element activity, often measured through methylation of CpG (5-mC) dinucleotides. While a dozen clinical syndromes are linked to defective imprinting, the extent this epigenetic phenomenon is linked to phenotypic variation and disease susceptibility remains undetermined. We show long-read HiFi genome sequencing for single-molecular profiling of 5-mC, together with pedigree-based phasing in early developmental tissue, provides critical insight into previously uncharted loci in the human genome. Using this approach in 75 samples from 25 trios, we develop a 10-fold enhanced map of human imprinting during development. The majority of POE was maternal (90%) with germ cell hypermethylation was confirmed at most loci (72%) showing signature of paternal imprinting. Integrating summary statistics from population GWAS finds enrichment of common (birthweight) and rare (congenital anomalies) disease loci in newly identified imprinting regions. Accessing pedigree-based rare disease cohorts, we show preponderance of paternal inheritance of pathogenic variants mapping to autosomal dominant OMIM genes with a maternal POE-bin identifying two genes (*BNC2*, *DNMT1*) as novel candidate imprinting disorder loci. Our enhanced human map of POE of 5-mC significantly extends the current “imprintome” and uncovers previously underappreciated genes and variants that appear crucial for human development and disease.

## Main

Genomic imprinting is thought to be mainly established in spermatocytes and oocytes and passed on to offspring and maintained in the early embryo before being erased in later developmental stages. Mechanistically, genomic imprinting involves differential inactivation of regulatory elements, often through 5-mC as the proximal regulatory mark for unequal, or imprinted, expression of genes in *cis*. Current estimates of the number of human imprinted genes range from several dozen to a few hundred, depending on the method and extent of experimental analyses employed^1^.

Imprinted genes are essential in regulating placental function, fetal development and growth, with at least a dozen known rare diseases caused by defects in genomic imprinting^2^. The genomic locations for these diseases cluster in imprinting control regions (ICRs)^3^ on chromosomes 7, 8, 11, 14, 15 and 20. While these diseases exhibit heterogenous clinical presentation, they share not only features associated with developmental delay and growth disturbances but also placental phenotypes. The unique regulation of these loci leads to unusual spectrum of mutational mechanisms as compared to typical Mendelian diseases^4^ including structural variation or 5-mC defects at ICRs, uniparental disomy as well as point mutations in imprinted genes (manifesting if inherited from one parent). Similarly, these imprinted genes can contribute to complex trait variation^5–7^.

Optimal approach for detecting POE of 5-mC relies on separately assessing functional output from each allele assigned as maternal or paternal and subsequently allele-specific methylation (ASM) assessment. However, direct mapping of ASM and the untangling of complex imprinted regions at the intragenic scale have presented significant technical challenges. Methods such as whole genome bisulfite sequencing (WGBS)^8,9^ and methylation-sensitive restriction enzymes (MSRE)^10^ have been used to identify differentially methylated regions (DMRs), including those of known imprinted regions, but often these methods focus on allelic contrast that does not distinguish POE. Phasing of WGBS into parental haplotypes is limited by short read lengths, and in typical paired-end bisulfite sequencing only ∼7% of reads are informative for allelic effects^11^. Large-scale studies by WGBS in mature peripheral blood tissue as opposed to early developmental have shown limited discovery of novel imprinting regions^12^.

In contrast to WGBS, current 3^rd^ generation platforms, such as PacBio’s Single Molecule, Real-Time (SMRT) HiFi-GS technology, generates 5-mC profiles genome-wide from standard sequencing libraries of long reads (∼16 kb) increasing phasing efficiently over tenfold as compared to WGBS. This improved haploid resolution of 5-mC detection is critical for POE discovery. We recently released a comprehensive 5-mC HiFi-GS data set from 276 rare disease patients for parallel variant and 5-mC calling^13^. We first validated the method through comparisons with WGBS and showed high concordance in 5-mC. Then, we used the HiFi-GS data set to identify >50,000 rare hypermethylation events that were to a large extent (80%) allele-specific and predicted to cause loss of regulatory element activity (LREA). We exemplified the power of HiFi-GS in unsolved rare disease cases by identifying a previously unknown, disease causing LREA event^13^. Consequently, it is likely that similarly to understanding unconventional genetic variants in rare disease cases, HiFi-GS will allow detection of genomic imprinting at a scale not appreciated before with potential for discoveries of new imprinting disorders.

Here, we used the established 5-mC HiFi-GS platform and sequenced early (6-8 weeks gestation) developmental fetal (chorionic villi) and parental peripheral blood tissue at high depth, respectively, for pedigree-based phasing of HiFi-GS reads. By applying a genome-wide segmentation approach, we identify 52,786 autosomal CpGs in 5852 bins showing POE of CpG methylation (POE-me) of which 60% have previously not been linked to or suggestive of imprinting. We perform systematic validation applying 5-mC HiFi-GS on germ cells, as well as blood biospecimens retrieved across development. Finally, accessing GWAS summary statistics from large biobanks testing genetic impact of common and rare disease traits along with trio-based rare disease cohorts, we show links between our imprinting regions and POE on human disease risk.

## Results

### 5-mC HiFi-GS of Early Developmental Tissue

We applied 5-mC HiFi-GS including simultaneous long-read genome and methylome profiling of 19 complete trios consisting of fetal chorionic villi (**Supplementary Table 1**) and parental blood to an average depth of 39X phasing each of the ∼27 million autosomal CpGs into maternal and paternal haplotypes corresponding to 98% of CpGs with accurate phasing. We performed genome-wide segmentation of each pedigree-phased fetal sample into ASM regions requiring complete consistency of paternal and maternal transmission across all samples as outlined in detail in **Supplementary Figure 1** – ensuring observed ASM patterns robustly exclude effects from DNA sequence variants, commonly known as *cis* methylation quantitative trait loci (*cis*-meQTLs). Using this approach, we identified 52,786 autosomal CpGs mapping to 5852 POE-me bins (**Supplementary Table 2; Figure 1A**) of which 10% were paternally hypermethylated (N=591 paternal POE-me bins). We noted lack of POE-me bins on short arms of acrocentric chromosomes linked to their complex, repetitive sequences resulting in low coverage (**Supplementary Figure 2A**). No impact of aneuploidy was observed for the 5852 POE-me bins (**Supplementary Figure 2B**).

**Figure 1.**
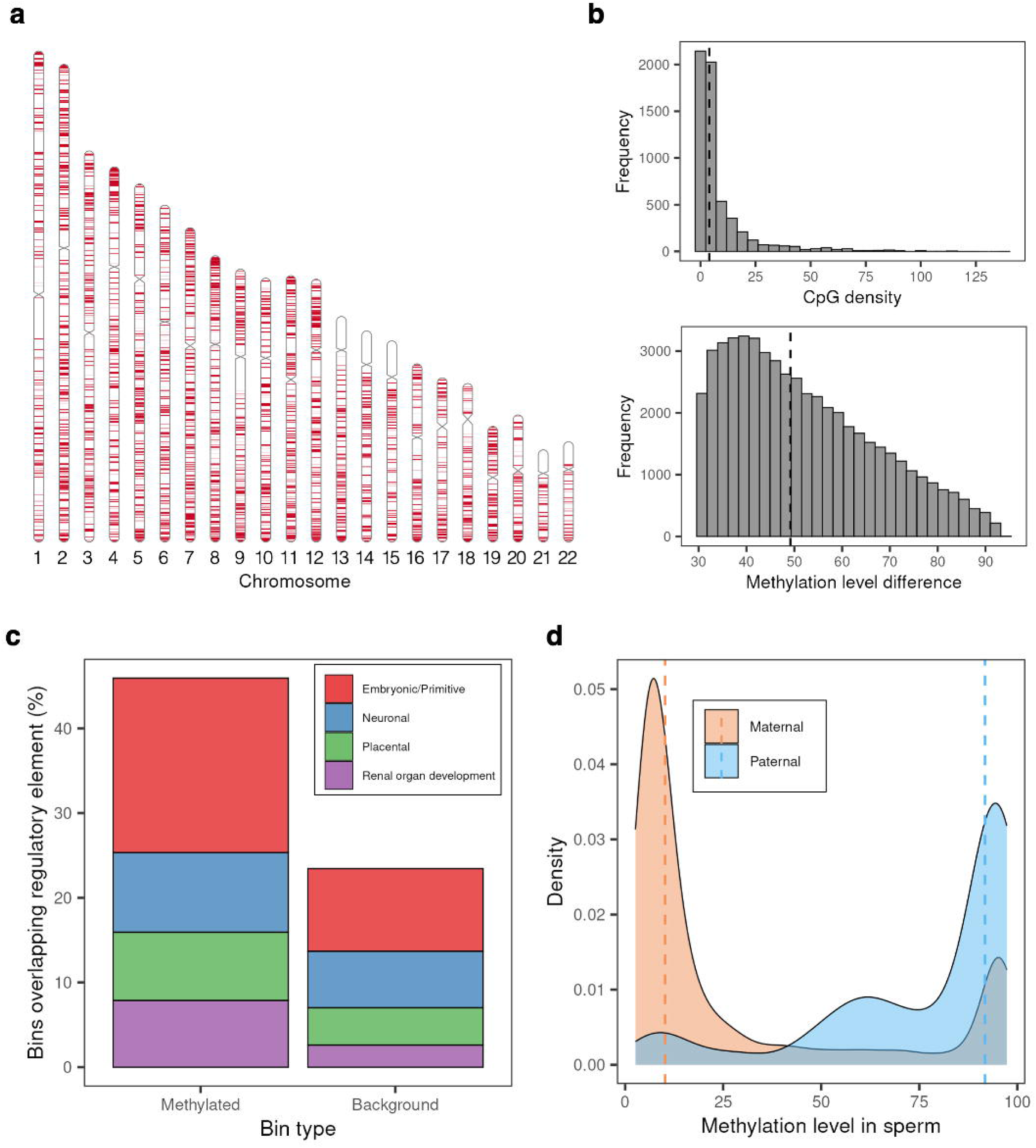
**A.** Map of POE-me CpGs (red) across chromosomes. **B.** Histogram showing CpG density (top) and methylation difference (bottom) across all POE-me bins. **C.** Fraction of POE-me and background bins mapping to indicated regulatory element. **D.** Methylation level in sperm for maternal (orange) and paternal (blue) POE-me bins.

The median CpG density and methylation level difference in the POE-bins was 4 and 49%, respectively (**Figure 1B**). Higher CpG density correlated with larger methylation differences in each POE-me bin (**Supplementary Figure 3**). In total of 1674 of the 5852 (29%) POE-me bins mapped within 20kb of the transcription start site (TSS) of a known gene as determined by GREAT (**Supplementary Figure 4**). Restricting to POE-me bins mapping within 20Kb of protein coding regions (1021 genes) uncovered top five GeneOntology (GO) term enrichment for neuronal system (log_10_ q value=-8.55), chemical synaptic transmission (log_10_ q value=-8.55), behavior (log_10_ q value=-8.39), neuron projection development (log_10_ q value=-6.88) and sensory organ development (log_10_ q value=-5.34) (**Supplementary Figure 5A**). Enrichment analysis for cell type signatures revealed neurons and glial cells as most significant (**Supplementary Figure 5B**).

Overlap using comprehensive GENCODE V45 annotation including both coding and non-coding transcripts revealed 4,915 (84.0%) of the 5,852 POE-me bins mapping within 20kb of a gene (**Supplementary Figure 6**). POE-me bins mapped more often to regulatory elements specific to cells of the following origin: embryonic/primitive (2.1-fold, P=1.39 × 10^−59^, Chi^2^ test), renal organ development (3.0-fold, P=2.14 × 10^−37^, Chi^2^ test), placental (1.8-fold, P=5.58 × 10^− 16^, Chi^2^ test), and neuronal (1.4-fold, P=5.20 × 10^−8^, Chi^2^ test), respectively as compared to background control regions (**Figure 1C**).

### Validation of Early Developmental POE-me Pattern

We performed several validation analyses to ensure robustness and accuracy of the approach to discover novel imprinting. First, we consolidated a list of well-established loci defined as discovered by at least two independent human studies^12,14–16^ corresponding to 116 regions of which 102 (88%) regions were among our final set of POE-me bins (**Supplementary Table 3**). We manually curated the 14 regions not replicating, noticing 11 of these were identified in our ASM mapping pipeline but not reaching our final stringent criteria for putative imprinting (**Supplementary Table 3).** Indeed, only three regions mapping to chromosome 15 were not showing evidence of imprinting likely being blood-specific^12,15^ with prior placental studies also missing these signatures^17,18^. Next, we assembled a replication cohort of six additional trios consisting of fetal chorionic villi and parental blood and similarly performed high-depth 5-mC HiFi-GS followed by complete phasing to obtain haploid-resolved methylation profiles (**Supplementary Figure 1**). We required POE-me CpGs to be covered in all samples (N=22,554) and tested whether the observed proportions of methylated reads were significantly different in paternal vs. maternal haplotypes using Fisher’s exact test. Using strict Bonferroni corrected P-value (0.05/22554=2.242 × 10^−6^) and the same direction of effect as the discovery set, we noted an 85% replication rate at single CpG resolution. Overall, the replicated correlation of ASM differences across all 22,554 CpGs was high (Spearman R=0.88, **Supplementary Figure 7)**.

We also incorporated candidate^17,18^ and confirmed loci reported by others and noted that out of our 5852 POE-me bins, 60% (N=3501) are identified for the first time, 21% (N=1224) have been proposed but not previously parentally confirmed and 19% (N=1127) have been reported earlier by at least two studies (**Supplementary Table 4).** Focusing specifically on suggested imprinting loci^17,18^ in placenta tissue detected using ASM-based approaches, we however noted a higher replication rate across the two studies (∼45%) than across either of ASM studies and our POE-me bins (∼30%; **Supplementary Table 5)**. We hypothesized that this increase in replication rate across the ASM-based imprinting studies may be inflated by genetically regulated loci. To test this, we calculated mean haplotype-resolved (maternal vs paternal) methylation level for the candidate regions identified by others (**Supplementary Table 5)** using our fetal discovery samples. We then examined the interindividual variation in the directionality of ASM. Notably, we observed significant differences in consistency of ASM based on the approach used for the discovery of imprinting, with ASM regions validated by our POE-me bins exhibiting largely unidirectional patterns (**Supplementary Figure 8**).

Altogether, these results show superior value of haplotype-based methods for discovery of imprinting signatures leading to more than 400% increase in the confirmed and validated imprinted loci in humans.

### Genetic vs Non-Genetic Effects of Allelic DNA Methylation

We compared the overall distribution of common allelic effects in methylation to identified POE-me in the fetal samples (n=19). Across all samples we identified nearly 19 million (18,391,349) 200bp bins with recurrent (observed in at least 5 individuals) ≥2-fold differences in allelic methylation (minimum 2 CpGs per bin). On average we observed 967,966 variable bins per individual of which 1,899 (0.2%) overlapped known and 6,074 (0.6%) overlapped novel POE-me bins. POE-me bins show strong maternal hypermethylation bias (80 – 92% maternal) as shown above whereas among the rest of allelic methylation variation (99.2%), the relative parental bias is almost equal (55% maternal hypermethylation bias). This suggests that among the “non-POE” allelic methylation the heritable effects of allelic methylation^13^ may be predominant with minor residual POE not captured by our strict approach for finding novel imprinting. The transmission of allelic methylation can be only partially studied in our trios given the overall different methylation patterns in parental blood vs. fetal tissue. However, among the loci where parental blood allelic methylation showed ≥2-fold differences the relatively hypermethylated haplotype was shared among parent -and offspring significantly more frequently (P<0.000001, Chi^2^ test) among “non-POE” bins (62,7% of 1,16M bins) as compared to known POE bins (54.5% of 1,069 bins) suggesting stronger haplotype specific heritability for the former. Altogether, like earlier observations^19^, allelic variation is very common and enriched for genetic effects with consistent POE accounting for less than 1% variation overall^20^.

### POE-me Across Tissue and Development

Early developmental tissue is commonly utilized for characterizing human imprinting as robustly validated genes showing POE are often linked to placental function and fetal growth and prior efforts have found only limited number of genes showing POE in adult tissues^12,21^. However, systematic comparisons of POE signatures across large tissue cohorts using the same analytical framework have not been done previously. To this end, we assembled a pedigree-based cohort comprised of peripheral blood samples from a large pediatric population (N=127) using 5-mC HiFi-GS at high depth (**Supplementary Figure 1**). We required POE-me CpGs to be covered in >25 samples (N=47,803). We found 10% (N=4979) of the POE-me CpGs showing significant (Bonferroni corrected P 0.05/47,803=1.05 × 10^−6^, Fisher’s exact test) differences in the same direction as our CpGs identified in early developmental tissue (**Supplementary Table 6**). With the notion that early developmental signatures of POE fade in somatic tissues, we allowed a less conservative threshold of significance for estimation of putative imprinting signatures in blood. This resulted in 18% (N=8734) of the POE-me CpGs showed significant (P<0.05, Fisher’s exact test) differences in the same direction as our CpGs identified in early developmental tissue.

### Integration of Germ Cell HiFi-GS

Genomic imprinting is believed to be established before fertilization, during the development of the male and female gametes. While non-canonical imprinting, manifesting as post-fertilization POE has been proposed, recent reports show that this phenomenon is not conserved in humans^22^. To gain insight into this, we generated 5-mC HiFi-GS at high depth on male germ cells from a cohort of 23 samples derived from a sperm bank of anonymous donors. We noted drastic differences in sperm methylation level of POE-me CpGs being identified as maternal vs. paternal (**Supplementary Table 7**). Specifically, 72% (N=1992) of POE-me CpGs showing paternal imprinting in chorionic villi were hypermethylated (defined as >50% methylation level) in sperm samples corresponding to 85% of the POE-me bins (**Figure 1D**). On the other hand, similar assessment of POE-me CpGs showing maternal imprinting in chorionic villi revealed only 7% (N=3681) being hypermethylated in sperm with 90% of maternal POE-me CpGs being hypomethylated (defined as <25% methylation level) in sperm samples (**Figure 1D**). While these results point towards majority of paternal imprinting being established prior to fertilization, we investigated the subset of paternal POE-me bins showing complete hypomethylation in sperm (N=40 POE-me bins) restricting to those with five or more CpGs (N=22/40 POE-me bins; **Supplementary Table 7**). Interestingly, 20 out of those 22 paternal POE-me bins (**Supplementary Table 8)** were mapping near (within 50kb) a maternal POE-me bins potentially indicating evidence of bipolar dominance effects as described before^23,24^ however with compensatory regulatory effect on opposite allele occurring post-fertilization.

### POE-me, Common Genetic Variation and Complex Traits

Common genetic variants mapping to genomic imprinting regions can have different phenotypic effect based on parental origin as shown for fetal growth-related traits^7,25^, cardiovascular^26^ and metabolic traits^27^ etc. To test this, we first assessed overlap of our POE-me bins with GWAS summary results of birth weight from a large cohort study^25,28^ taking both parental and fetal genomes into account. We identified a significant enrichment of birthweight associated variants overlapping our POE-me bins compared to background bins (**Figure 2A**). We then tested for differences in birth weight across GWAS variants resolved for parental inheritance in two trio-based cohorts with available birthweight data (DDD: N = 1,578; GA4K: N = 235; **Methods**). In the DDD cohort, we observed a greater effect for maternal versus paternal inheritance of POE-overlapping variants compared to background variants. This effect increased as a function of GWAS P-value (**Figure 2B)**. Although limited by available birth weight measurements in GA4K, for GWAS variants with at least nominal significance (GWAS P<0.05) effect on birth weight in DDD that could also be tested in GA4K (N = 11 variants) we observed high concordance in effect direction (9/11, 82%) and correlation of effect magnitude (Pearson’s correlation r= 0.52; P-value = 0.04). In the DDD cohort, we further assessed differences in the proportion of probands with GWAS POE-me or background-overlapping variants with respect to birth weight outlier status. We observed an increasingly larger fraction of probands with POE-me overlapping variant(s) (filtered for parental inheritance, e.g. maternal POE-me bin with paternal inheritance) as a function of outlier Z-score (**Figure 2C**). We observed no differences in the proportion of probands with background variant(s) as a function of birth weight outlier status. We then selected probands with POE-me overlapping putatively methylated alleles (e.g. maternal POE-me bin with maternal inheritance) and observed no differences in the proportion of probands with POE-overlapping variant(s) as a function of outlier status (**Figure 2D**).

**Figure 2.**
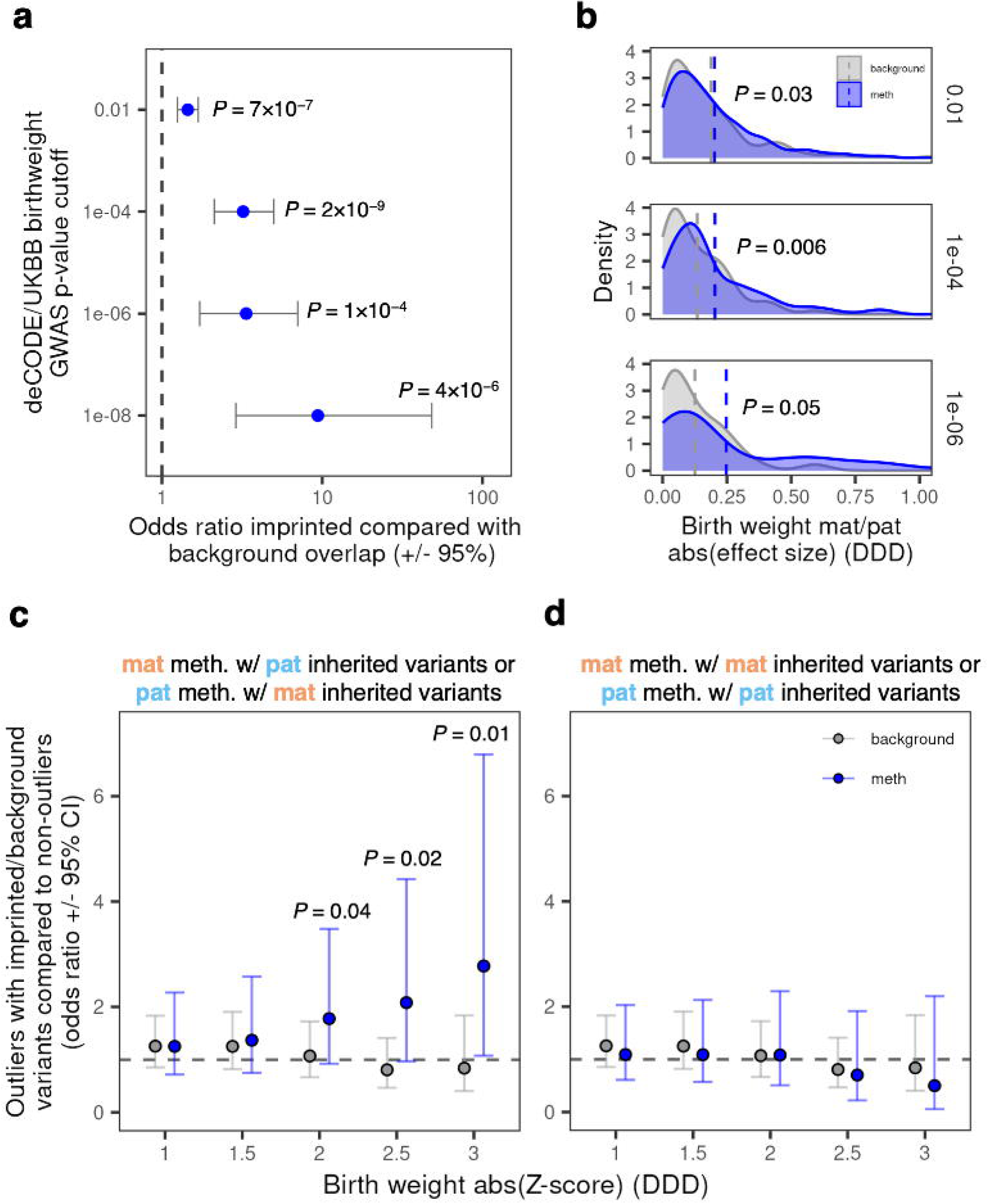
**A.** Odds ratios comparing POE-me and background bins overlapping birth weight GWAS hits, stratified by GWAS P-value. **B.** Logistic regression coefficients comparing effect on birthweight in DDD probands of paternally or maternally inherited variants for POE-me (blue) and background (gray) bins, stratified by birthweight GWAS P-value. **C.** Odds ratios comparing the proportion of probands with POE-me (blue) or background (gray) GWAS overlapping variant as a function of birth weight Z-score and for POE-me parental inheritance indicating a non-methylated allele. **D.** As in (**C.**) but for POE-me parental inheritance indicative a methylated allele.

Restricting to those POE-me bins near a birth weight GWAS variant at P<1E-8 (N=72, **Supplementary Table 9**), we noted different features as compared to our total set of POE-me bins (N=5852) as follows: 1) larger proportion of the POE-me bins being paternally inherited (49% vs 10%, Chi^2^ P=8.58 × 10^−27^), 2) larger proportion mapping to a regulatory element (78% vs 46% Chi^2^ P=1.49 × 10^−7^), and 3) larger proportion showing overlapping trend in blood tissue at nominal p<0.05 (37% vs 24% Chi^2^ P=0.046). No enrichment of genetic variants near our POE-me bins were observed for birth length (**Supplementary Figure 9**).

Next, we integrated GWAS results from a recent study using large biobanks arguing that POE can be computationally inferred without the use of parental genomes for estimation of phenotypic effects. While only three loci were reported by Hofmeister et al with additive effects in their genome-wide analysis, we noted that two out of the three (67%) loci were mapping to a novel POE-me bin including a novel paternal POE-me bin near *TERT* associated with relative telomere length (rs2735940) and a previously unreported maternal POE-me bin at the known *KCNQ1* locus associated with standing height (rs143840904). While we don’t have access to telomere length in our pedigree-based GA4K cohort, we tested for POE in height for variants included in a recent large-scale saturation GWAS^29^ that also overlapped a POE-me bin and compared with a matched background set. Among probands in GA4K where variant inheritance could be directly resolved, we observed a larger effect for differences in height for maternal versus paternal inheritance in POE-me overlapping variants compared to background. This effect increased as a function of GWAS P-value (**Supplementary Figure 10**).

### POE-me, Rare Genetic Variation and Complex Traits

Large biobanks with high-resolution genotype or sequencing data are allowing investigation with progressively rarer alleles. We utilized a filtered set of GWAS summary statistics from the Finngen R12 release^28^ comprising 21,311,644 genetic variants across 500,348 individuals (**Methods**). By integrating significant GWAS hits (P<1 × 10^−8^) with our POE-me bins, we identified a significant enrichment of genetic risk variants associated with phenotypes grouped as congenital malformations, deformations and chromosomal abnormalities mapping in or near (within 10kb) our POE-me bins compared to background bins (**Figure 3A**). Allele frequencies (AF) of significant variants ranged from 0.01%-50% and mapped near 49 unique POE-me bins (**Supplementary Table 10**). The top locus (rs572078678; P=1.97 × 10^−61^; beta=43; AF=0.0008) from the enrichment analysis was driven by rare variants from GWAS of congenital malformations of gallbladder, bile ducts and liver (315 cases and 498691 controls) (**Figure 3B**) mapping to maternal POE-me bins overlapping the *DNMT1* promoter. While *DNMT1* is known to have maternal hypermethylation in its promoter^14,15^ across tissues including blood as replicated here, we show here that the imprinting region is larger than previously estimated spanning ∼7kb (**Figure 3C**). Interestingly, the extended POE-me region at the *DNMT1* locus is not showing evidence of POE-me in blood tissues (**Supplementary Table 6**). Long-read, isoform expression profiling in four fetal chorionic villi samples show almost exclusive paternal expression of the known somatic isoform (s-*DNMT1*) with start site near our maternal POE-me bins as well similar striking, paternal expression of a novel, shorter isoform slightly further downstream (**Supplementary Table 11**; **Supplementary Figure 11**). While we also detect strong expression of the oocyte specific variant (known as the *DNMT1o* isoform), this isoform is not showing evidence of POE of expression. Additionally, a smaller paternal POE-me bin detected in our analysis correlated with a novel, maternally expressed *DNMT1* isoform further downstream (**Supplementary Figure 11**).

**Figure 3.**
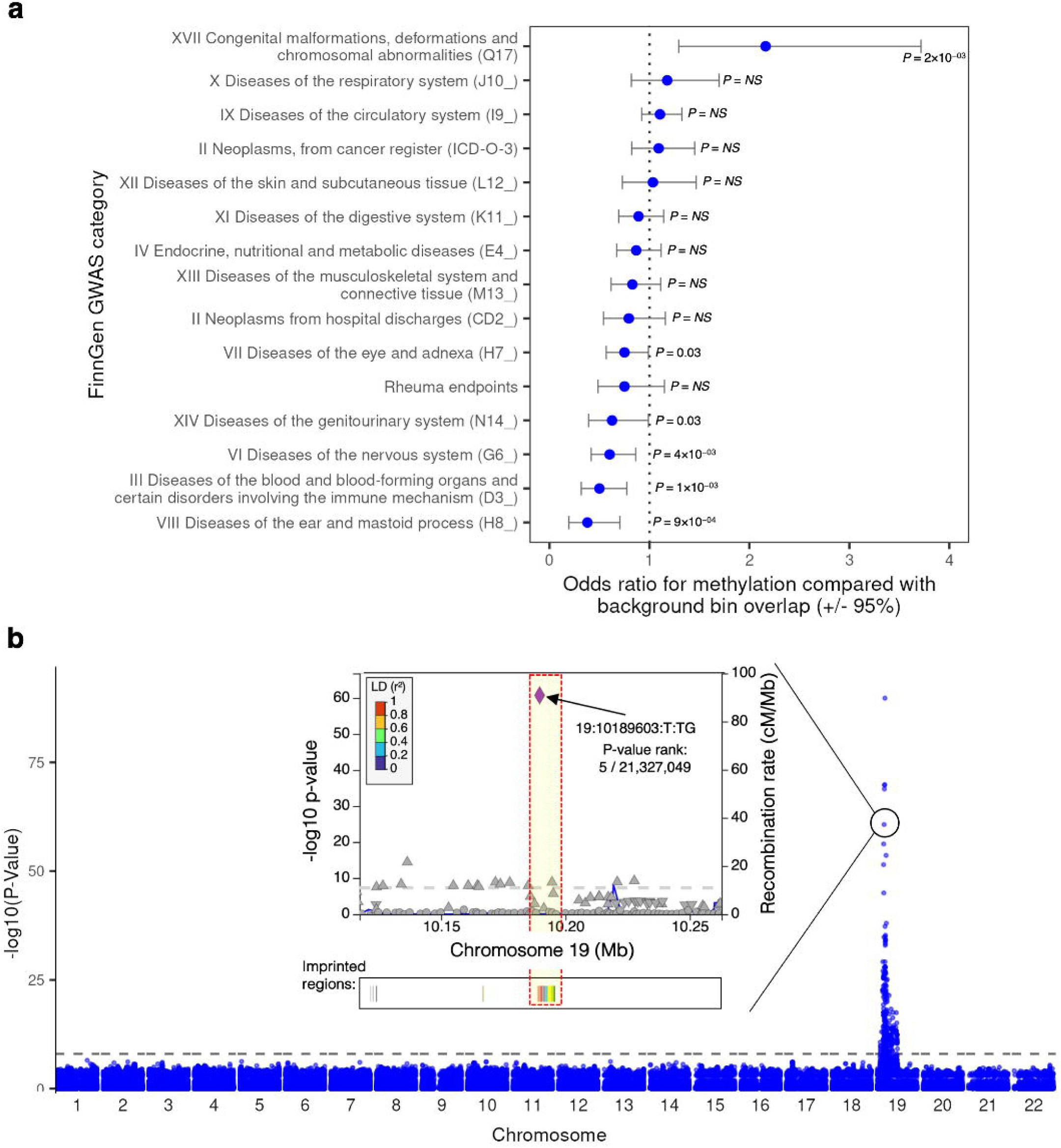
**A.** Odds ratios comparing POE-me and background bins overlapping GWAS hits for indicated FinnGen phenotype categories. **B.** Manhattan plot with inset showing overlap between POE-me bins and a FinnGen GWAS hit for “congenital malformations of gallbladder, bile ducts and liver”.

### POE-me bins Inform Rare Disease Diagnoses

With the notion that point mutations in imprinted genes can cause congenital disease if it affects the expressed allele^30^, we sought to explore this phenomenon for genes mapping to our POE-me bins. For this purpose, we utilized full phenotypic and sequencing access to the DDD^31^ and GA4K^32^ family-based rare disease cohorts of 9859 and 2057 trios, respectively with the DDD representing the discovery set and GA4K replication due to its smaller size. In addition, we benefit from access to summary level variant transmission data from our clinical genetics and genomics lab repository (CGGL) for further replication. We extracted rare (GnomAD AF<1×10^−5^) coding missense or loss of function (LOF) variants mapping to genes within 20kb of a POE-me bin. We further classified overlapped genes based on OMIM mode of inheritance and missense variants as benign or pathogenic using the AlphaMissense score^33^. We observed a significant enrichment of genes (**Supplementary Table 12**) with a POE-bin being classified as autosomal dominant (AD) compared to a control gene set indicative of monoallelic expression and this association was strengthened with increasing POE-me bin size (**Figure 4A**) and replicated using the DDG2P classification (**Supplementary Figure 12; Supplementary Table 12**). The association of AD inheritance for genes with a POE-me bin was maintained when known imprinted genes were removed (**Supplementary Figure 13**).

**Figure 4.**
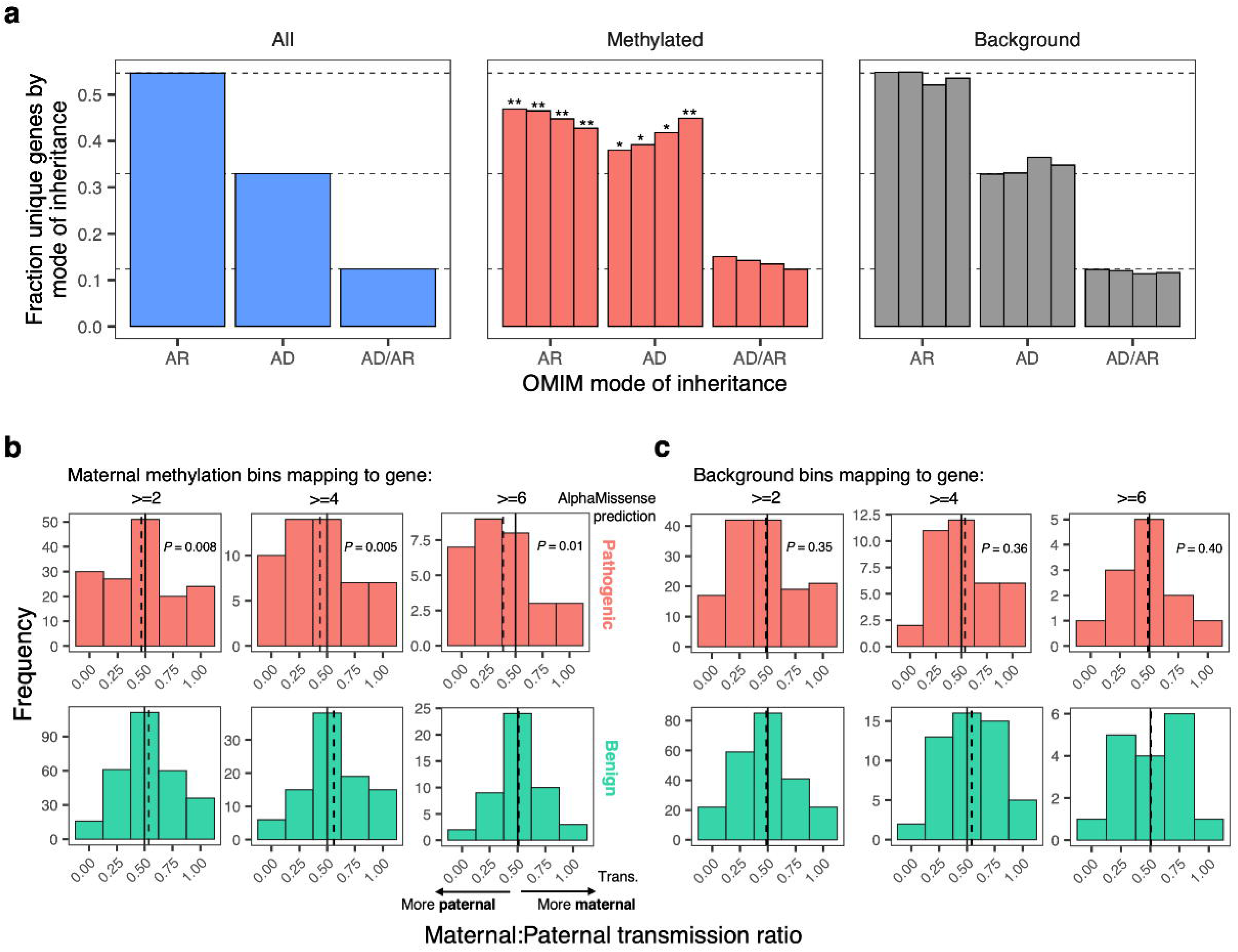
**A.** Fraction of genes for each indicated mode of inheritance in OMIM (blue), OMIM genes overlapping POE-me bins (red), and OMIM genes overlapping background bins (blue). POE-me and background bins are stratified by bin size (from left to right: any bin; >=100 bp; >=250 bp; >=500 bp). **B.** Maternal-to-paternal transmission ratios for predicated pathogenic (top; red) and benign (bottom; green) variants in genes overlapping maternal POE-me bins. Results are stratified by number of bins mapping to a gene (from left to right: >=2 bins; >=4 bins; >=6 bins). **C.** Maternal-to-paternal transmission ratios for predicated pathogenic (top; red) and benign (bottom; green) variants in genes overlapping background bins. Results are stratified by number of bins mapping to a gene (from left to right: >=2 bins;>=4 bins; >=6 bins).

Next, we focused on genes possessing maternal POE-me bins specifically and contrasted inheritance of rare variants classified as benign vs pathogenic. We found a significant difference of the ratio of maternal:paternal inheritance of variants classified as pathogenic vs benign. While the latter showed a median ratio of inheritance as 50:50, pathogenic variants had an expected skewed ratio towards paternal inheritance, and this association was magnified when analysis was restricted to genes with large maternal POE-me bins (**Figure 4B**; P<0.01). Importantly, this trend was not observed when the analysis was repeated for genes not associated with a maternal POE-me bin (**Figure 4C**). While the low number of paternal POE-me bins prevented robust statistical analysis, regardless, the same trend was observed for the ratio of maternal:paternal inheritance of pathogenic variants mapping to genes with a paternal POE-bin showing preferential maternal inheritance (**Supplementary Figure 14**). We then focused on a subset of genes with maternal POE-bins with deviation from the 50:50 maternal:paternal inheritance ratio of pathogenic variants requiring 75% or more of pathogenic variants per gene to be paternally inherited (N=34 genes). Restricting further to 1) requiring OMIM classification (N=10 genes) with 2) AD inheritance pattern resulted in five candidates we put forward for replication analysis in GA4K and CGGL, respectively: *BNC2*, *DNMT1*, *KIF1A*, *PDE3A* and *TENM4* (**Table 1**). *BNC2* had consistent inheritance pattern (paternal) for pathogenic variants across all three trio-resolved cohorts and while *DNMT1* could only be tested in DDD, it remains a strong candidate for future analysis given its link to congenital malformations (**Figure 3**).

**Table 1:** Candidate imprinting disease locus.

| **Gene** | **maternal**<br>**POE-me**<br>**bins (N)** | **path**<br>**variants**<br>**– DDD**<br>**(N)** | **paternal**<br>**inheritance**<br>**DDD (%)** | **path**<br>**variants**<br>**– GA4K**<br>**(N)** | **paternal**<br>**inheritance**<br>**GA4K (%)** | **path**<br>**variants**<br>**– CGGL**<br>**(N)** | **paternal**<br>**inheritance**<br>**CGGL (%)** |
| --- | --- | --- | --- | --- | --- | --- | --- |
| *BNC2* | 3 | 4 | 75 | 1 | 100 | 2 | 100 |
| *DNMT1* | 10 | 2 | 100 | 0 | NA | 0 | NA |
| *KIF1A* | 7 | 3 | 100 | 14 | 72 | 2 | 50 |
| *PDE3A* | 7 | 3 | 100 | 0 | NA | 1 | 0 |
| *TENM4* | 3 | 2 | 100 | 10 | 80 | 4 | 50 |

### POE on Rare and Common Phenotypes at the BNC2 Locus

Mice lacking *BNC2* die within 24 h of birth with a cleft palate and abnormalities of craniofacial bones and tongue^34^ including smaller heads and high incidence of distal urethral defects^35^. In humans, rare variants in *BNC2* have been linked to Autosomal-Dominant Congenital Lower Urinary-Tract Obstruction (LUTO)^36^. In addition, GWAS have shown association of *BNC2* SNVs with Adolescent Idiopathic Scoliosis (AIS)^37^ and human height^29^ with top genetic variants mapping ∼6kb downstream of our POE-me bin (**Figure 5**). Here we use long-read, isoform expression profiling and identify an isoform originating from our POE-me bin (**Supplementary Figure 15**).

**Figure 5.**
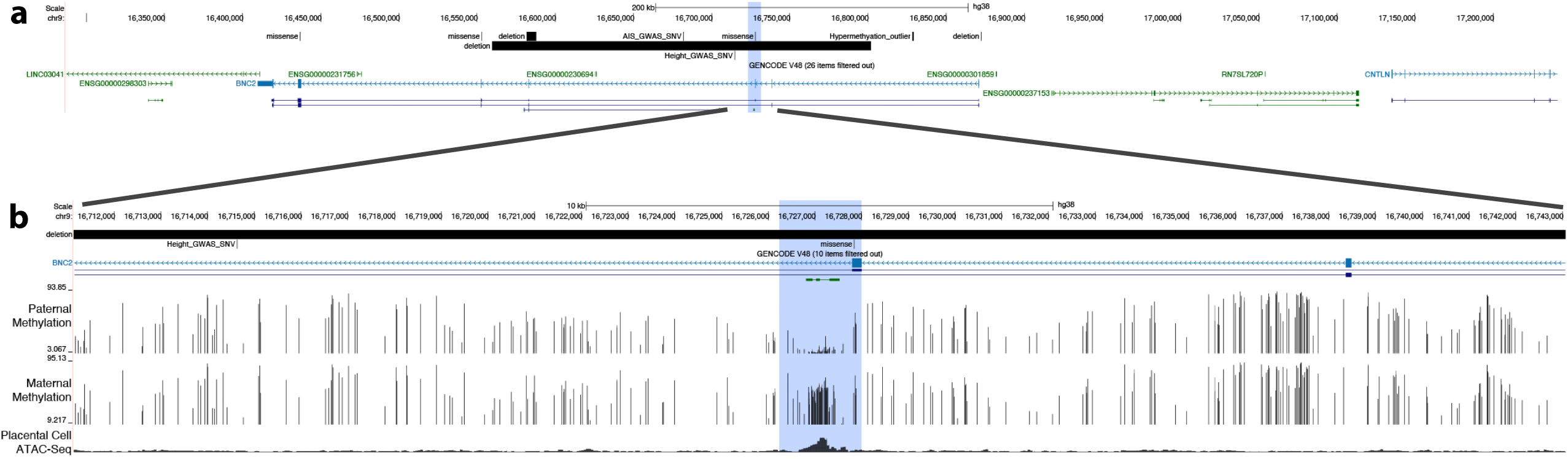
**A.** Genomics view of the *BNC2* locus highlighting the maternal POE-me bin (blue box) and assessed genetic variants, deletions and hypermethylation outlier indicated based on their size (black bars). **B.** Zoomed in view of the *BNC2* locus centered around the POE-me bin (blue box). Upper tracks shows paternal and maternal CpGs methylation (y-axis, %) measured by 5mC HiFi-GS and bottom trac shows open chromatin peas measured by ATAC-Seq in trophoblast stem (placental) cells.

We reviewed the three cases (all paternally inherited) from our systematic analysis across the GA4K and CGGL cohorts (**Table 1**) where we had detailed clinical information available (**Supplementary Table 13**). These cases remain unsolved and have been clinically signed out as non-diagnostic. Intriguingly, in line with *BNC2* animal modeling and human GWAS, one case had severe facial dysmorphism and skeletal alternations including high palate and early-onset scoliosis and another case had short stature and congenital heart defects (**Figure 5**). We expanded our search to our CGGL cytogenic databases identifying three additional cases with *BNC2* deletions (**Figure 5**). All three cases remain non-diagnostic and deletions were reported as a variant of unknown significance (VUS). While the inheritance remained unknown the clinical presentation was very similar to the cases with the *BNC2* missense mutations. Here, all three cases had facial dysmorphism, skeletal alterations and/or congenital heart defects (**Supplementary Table 14)**.

Next, we utilized an approach described earlier for rare hypermethylation outlier discovery in 5mC-HiFi-GS data^13^ (**Methods**) using a larger sample set including 687 probands with family members (total N = 1,112). To reduce the number of rare disease relevant candidates we focused on a subset of probands with both parents also having a high depth 5mC-HiFi-GS (n=108) and required that 200bp rare outlier tiles showed extreme deviation (z > 3) from parental methylation and could be described as *de novo* in the proband. We overlapped these *de novo* tiles with TSS of known OMIM genes (+/- 10kb). A total of 171 such tiles were seen (1.6 per proband). We further restricted to OMIM AD genes where at least two tiles colocalized to same TSS proximal region, since known disease causing hypermethylation invariably spans several hundred bp. Intriguingly, only one OMIM AD gene/sample pair remained including two tiles in 1st intron of *BNC2* with no other diagnostic finding found in the genome of this proband with the hypermethylation tiles in paternal haplotype. The region of hypermethylation was seen overlapping nearly fully constraint DNA element and open chromatin in developing kidney among other tissues (**Figure 5**). The phenotype of this proband included severe facial dysmorphism, skeletal alterations, congenital heart defects and early perinatal death (**Supplementary Table 15)**. We then scanned the additional 579 probands with 5mC-HiFi-GS and found a second case with exactly same hypermethylation tiles on paternal allele both with z>4 across the full region and a shared phenotype with index *de novo* methylation case similarly showing severe facial dysmorphism, skeletal alterations, congenital heart defects and early perinatal death (**Supplementary Table 15)**.

Finally, given *BNC2* variant association to standing height in large-scale saturation GWAS^29^, we assessed differences in height and rate of clinical short stature trajectories using time-resolved observation data available in our pedigree-resolved GA4K cohort. The height associated variant at the *BNC2* locus, rs1330304 (p=6.15 × 10^−82^, EUR), is located 11.5Kb downstream of our maternal POE-me bin (**Figure 5**). In GA4K, we observed overall decreased height for probands with a paternally inherited effect allele compared with maternal, as well as an increase in the rate of clinical incidence of short stature (**Supplementary Figure 16**) indicative of POE of height phenotypes.

## Discussion

Studies on genomic imprinting have been ongoing for several decades, with robust evidence of its relevance to important biological processes such as epigenetic regulation and developmental disorders like Prader-Willi Syndrome and Angelman Syndrome. Yet, the number of regions in the human genome that has been proven to be imprinted is rather scarce despite large-scale efforts^12^ in adult blood tissue. Much of the diagnostic utility in human imprinting disorders is also focused on the small subset of loci showing persistent POE in blood. Consequently, the known developmental “decay” of genomic imprinting has stimulated imprinting discovery to earlier developmental lineages, including placental tissue, also allowing linking of placental and fetal growth phenotypes to disruption of imprinting. However, in these studies technological or study design limitations of tracking parent-of-origin informed ASM has led to suboptimal recall and precision of novel human imprinting. Here, we leverage our unique collection of early developmental fetal chorionic villi (placental) samples with parent-of-origin phased haplotypes accurately generated by the HiFi long-read sequencing platform. We rigorously followed-up and validated the POE-me loci in independent fetal chorionic villi as well as germ cell DNA analyses. These efforts guided the generation of an enhanced map of the human imprintome extending known imprinting regions in their size as well as discovering thousands of new regions possessing imprinting signatures. A striking feature of these newly discovered regions is the predominance of maternal imprinting (90%) indicating a paternal bias in gene expression in this specific tissue. This observation is supported by early groundbreaking work showing the paternal genome being essential for normal development of extraembryonic tissue development^38^. Similarly, more recent work has presented evidence that paternally expressed genes indeed dominates not only in the placenta^39^ but also across other tissues^24^.

By further contrasting our map of new imprinted regions in extraembryonic tissue with similar map in adults blood tissue we show relatively poor overlap with less than 20% POE-me being detectable in blood. While we are unable to rule out overlap in in early embryonic tissues and/or neuronal tissue due to challenges is obtaining these sample types at scale, we speculate that dysfunctional placental imprinting can by itself cause disease phenotypes linked to e.g. brain and heart development^40^.

Understanding the potential role of the abundant imprinting during early development in human disease is challenging since most genomic cohort studies do not involve family members to allow firm parent-of-origin determination. Also, clinical genetics resources such as ClinVar do not systematically record parental origin of pathogenic alleles. Finally, outside rare disease diagnostics early life phenotypes are often uncaptured. Despite these limitations, we accessed accessible trios from Deciphering Developmental Disorders (DDD) and Genomic Answers for Kids (GA4K) consortia as well as available population cohorts and biobanks with relevant phenotypes and uncovered novel candidate parent-of-origin influences in rare and quantitative traits. The infrequent variant statistically linked to congenital anomalies of liver and biliary tract in *DNMT1* in a large biobank as well as common variants associated with birth weight replicated for POE in familial cohorts are all in novel confirmed imprinted regions from our study and not observable outside placenta. Similarly, *BNC2* known to cause autosomal dominant urogenital anomalies, shows evidence of variable paternally transmitted or paternal haplotype associated methylation variants with expanded phenotypic spectrum. This candidate imprinting disease locus is notable as a known AD condition, which are globally enriched among novel POE regions.

Altogether, our results suggest an underappreciated role of parental origin in genetic variation in disease. The vastly expanded catalog of developmental POE in human provides a roadmap for phenotypic and genotypic reclassification to investigate potential new imprinting diseases in patient and population cohorts. Moreover, the existence of hundreds of novel human loci under POE suggests that community resources reappraising the parental layer of variation would greatly enrich the overall interpretation of molecular changes in disease offsetting the higher cost of familial cohort establishment and parent-of-origin variant annotation.

## Online Methods

### Perinatal Study Cohort

Study participants were enrolled at the University of Kansas Health System Advanced Reproductive Medicine Clinic where they were diagnosed with recurrent pregnancy loss (RPL) using recommendations by the American Society for Reproductive Medicine Practice Committee^41^ with RPL defined as “spontaneous loss of two or more pregnancies.” For all patients known etiologies for miscarriage were ruled out before allowing participation in study including: Mullerian anomaly, polycystic ovary disease, thyroid disease, diabetes, coagulopathy, balanced translocation or structural rearrangement from sperm or oocyte contributor.

Miscarriage was diagnosed using American College of Obstetrician and Gynecologists Committee on Practice^42^, which defines miscarriage as “a nonviable, intrauterine pregnancy with either an empty gestational sac or a gestational sac containing an embryo or fetus without fetal cardiac activity within the first 12 6/7 weeks of gestation.” All study participants had transvaginal ultrasound to confirm diagnosis of miscarriage and had opted for dilation and curettage (D&C) procedure as a desired treatment option for surgical evacuation of miscarriage.

Product of conception (POC) (including extraembryonic placental and maternal decidual tissue) or embryonic tissue were collected during the D&C procedure. For POC samples, a section was carefully dissected by trained cytogenetics technicians to retrieve the chorionic villi and stored at 4°C in RPMI 1640 Medium (Irvine Scientific, 9161-500ML) until DNA isolation. Additional aliquot if available were stored at -80°C for RNA isolation.

Blood was obtained by trained phlebotomist from female participants and their partner via venipuncture and placed in lavender top tube with EDTA for anticoagulant.

Scavenged, left over semen samples were obtained from couples utilizing donor sperm in the *in vitro* fertilization process at the University of Kansas Health System Advanced Reproductive Medicine Clinic.

### Genomic Answer for Kids HiFi-GS Cohort

In total of 127 probands and their parents enrolled in the Genomic Answers for Kids (GA4K) program^13^ were included for blood replication of putative imprinting signatures. GA4K enrolls patients with a suspected rare disease referred to from multiple different specialties, with the largest proportion nominated by Clinical Genetics, followed by Neurology. A continuum of pediatric conditions is represented, ranging from congenital anomalies to more subtle neurological and neurobehavioral clinical presentations later in childhood.

Blood was obtained by trained phlebotomist from probands and their parents via venipuncture and placed in lavender top tube with EDTA for anticoagulant.

### DNA isolation and quality control

Perinatal (POC) tissue samples were subjected to DNA isolation using a CLIA/CAP protocol. In total of 30mg of tissue was removed from the sample and transferred to a sterile petri dish on ice. The tissue was then finely minced using a sterile scalpel and transferred into a 15mL conical tube. Next, 500µL of buffer (10mM of 1M Tris-HCl (Thermo Fisher, J22638AE), 10mM of 1M KCl (Quality Biological, 351-044-101), 10mM of 1M MgCl2 (Invitrogen, AM9530-G), 32µL/mL of 20% SDS (G Biosciences, 786-017), 2mM of 0.5M EDTA, pH 8.0 (Fisher Scientific, BP2482-100), 0.4M of 5M NaCl (Sigma, 71386-1L) and molecular-grade water) was added to the tissue and the tissue was agitated by pipette mixing. 40uL of 10mg/mL proteinase K solution was added to the tissue and the tissue was then incubated at 55°C overnight. After 12 hours, the sample was agitated by pipette mixing again to ensure it was thoroughly digested, and no tissue chunks remained in the sample. 225µL of 5M NaCl was added directly to the sample. The sample was mixed by vortexing and then centrifuged at 3500 rpm for 10 minutes at room temperature. The supernatant was transferred to a new 15mL conical tube without disturbing the protein pellet at the bottom of the original tube. 1µL of Pellet Paint (Millipore, 70748-125) was added to the supernatant to help visualize the DNA before the addition of 1.36mL of 100% molecular-grade ethanol. The sample was then thoroughly mixed by inversion, and the loose, blue, floating DNA pellet was spooled onto a sterile inoculating loop. The spooled DNA was gently rinsed of excess salt by dunking the inoculating loop in 1mL of 70% molecular-grade ethanol, being careful not to lose the DNA during the wash step. The inoculating loop was transferred to a 1mL barcoded Matrix tube (Fisher Scientific, 14-754-558) and the end of the loop was cut off at the neck of the tube. 300µL of TE buffer (Promega, V6231) was added to the tube and the sample was vortexed well and incubated at 55°C for 1 hour to dissolve the DNA. Once the isolation was complete and the DNA was dissolved, the DNA was assessed using 1µL of sample with a NanoDrop Spectrophotometer (Fisher Scientific, ND2000) to determine the A260/280 and A260/A230 ratios to verify the quality of the isolation. The DNA concentration and overall yield was quantified using a Qubit dsDNA Broad Range Assay Kit (Invitrogen, Q32853).

DNA isolation from EDTA blood was automated using the Chemagic 360 with a 1.8mL protocol (Revvity) following manufacturer’s guidelines. Once the isolation was complete and the DNA was dissolved, the DNA was assessed using 1µL of sample with a NanoDrop Spectrophotometer (Fisher Scientific, ND2000) to determine the A260/280 and A260/A230 ratios to verify the quality of the isolation. The DNA concentration and overall yield was quantified using a Qubit dsDNA Broad Range Assay Kit (Invitrogen, Q32853).

Semen samples were used for DNA isolation using a customized protocol implemented with the Qiagen DNeasy Blood & Tissue Kit (Qiagen 69506) and Masterpure Complete DNA and RNA Purification Kit (BioSearch Technologies, MC85200). Whole ejaculate samples were centrifuged at 600rcf for 5 minutes and supernatant was removed, and pellet was resuspended in 1ml of PBS. This step was repeated but following removal of the supernatant, the pellet was resuspended in 500ul of Lysis Buffer and pulse-vortexed for 5 minutes. Next, a mix of 486ul of nuclease-free water and 14ul of Proteinase K (15mg/ml stock from Qiagen DNeasy Blood Tissue Kit) were added to the lysis buffer and incubated at 56°C for 2 hours. The lysate was transferred to a 15-mL conical tube and 1 mL Buffer AL from the Qiagen DNeasy Blood and Tissue Kit was added and sample was mixed thoroughly by vortexing. Remaining steps were executed following manufacturer’s guidelines using the Qiagen DNeasy Blood Tissue Kit. Once the isolation was complete, samples were centrifuged in a SpeedVac for 85-90 minutes at 60°C, brought up to 200uL using buffer AE (Qiagen 69506), and stored overnight at 4°C. A modified protocol of Masterpure Complete DNA and RNA Purification Kit (BioSearch Technologies, MC85200) was then used to purify sperm DNA samples. Each sample’s volume was brought up to 300uL with 100uL of Tissue and Cell lysis solution (Biosearch Technologies, SS000401-D2). The protocol for F. Precipitation of total nucleic acids (for all biological samples) was followed using the sample. The DNA was dissolved, and the DNA was assessed using 1µL of sample with a NanoDrop Spectrophotometer (Fisher Scientific, ND2000) to determine the A260/280 and A260/A230 ratios to verify the quality of the isolation. The DNA concentration and overall yield was quantified using a Qubit dsDNA Broad Range Assay Kit (Invitrogen, Q32853).

### Aneuploidy Analysis

A subset of the POC DNA samples (**Supplementary Table 1**) were assessed for aneuploidy using SNP-microarray (ThemoFisher, Waltham, MA) performed in accordance with manufacturer guidelines and analyzed according to American College of Medical Genetics Guidelines^43^. Specifically, ThermoFisher Scientific CytoScanTM HD Platform was used including The ThermoFisher Scientific CytoScanTM HD CN+SNP microarray which is a targeted and whole genome array designed and manufactured by ThermoFisher Scientific. The CytoScanTM HD microarray contains ∼2,696,550 markers designed using human genome build GRCh37 (hg19). The chip has 1,953,246 non-polymorphic and 743,304 single nucleotide polymorphism (SNP) markers. The overall chip resolutions are as follows: 1 marker/880 bases for intragenic genes, 1 marker/1737 bases for intergenic genes and 1 marker/1148 bases for overall gene and non-gene backbone.

### PacBio HiFi long-read genome sequencing and analysis

A total of ∼5 ug of DNA per sample was sheared to a target size of 15-18 kb using the Diagenode Megaruptor 3 (Diagenode, Liege, Belgium). SMRTbell libraries were prepared with the SMRTbell prep kit 3.0 (102-182-700, Pacific Biosciences, Menlo Park, CA) following the manufacturer’s standard protocol (102-166-600) with some modifications as follows: Fragments longer than 10 kb were selected using the Sage Science PippinHT (Sage Science, Beverly, MA). Libraries were sequenced on the Revio System using the Revio Polymerase Kit (102-739-100) with 24 hr movies/SMRT cell. Samples were sequenced to a target of >25.8X coverage.

HiFi circular consensus reads using Google DeepConsensus and 5mC methylation base modification at CpG motifs, adding base modification (“MM”) and base modification probability (“ML”) BAM tags calls, were generated directly on-instrument using the PacBio Revio system. HiFi read mapping, variant calling, and genome assembly were performed using a WDL workflow (https://github.com/PacificBiosciences/wdl-common). HiFi reads were mapped to GRCh38 (GCA_000001405.15) with pbmm2 v1.10.0 (https://github.com/PacificBiosciences/pbmm2). Structural variants were called with pbsv v2.9.0 (https://github.com/PacificBiosciences/pbsv) with “--hifi --tandem-repeats human_GRCh38_no_alt_analysis_set.trf.bed” options to pbsv discover and “--hifi -m 20” options to pbsv call. Small variants were called with DeepVariant v1.5.0 following DeepVariant best practices for PacBio reads (https://github.com/google/deepvariant/blob/r1.3/docs/deepvariant-pacbio-model-case-study.md) and phased with WhatsHap v1.4. Pedigree phasing was done for the fetal samples using parental sequencing data, otherwise local phasing was used. Phase haplotype tags (“HP”) were added to the aligned BAM by WhatsHap. Pileup-based consensus methylation sites and probabilities were generated by the script “aligned_bam_to_cpg_scores.py” from pb-CpG-tools v2.3.2 (https://github.com/PacificBiosciences/pb-CpG-tools/) with the “-q 1 -m denovo -p model -c 10” options. Only CpGs covered at >=10x were considered for further analysis.

Aneuploidy analyses are performed using bioinformatic output reports generated for structural variant (pbSV, sawfish) break ends report of and HiFiCNV report for copy number variant (CNV).

Variant interpretation is performed using the 2015 ACMG Standards and guidelines for interpreting sequence variants^44^ and the 2020 ACMG Technical standards for interpreting and reporting constitutional copy-number variants^45^.

### Identification of novel imprinted regions

The segmentation workflow implemented in methbat (version methbat 0.12.0-ddc67bb; https://github.com/PacificBiosciences/MethBat) was run with the haplotype segmentation enabled using default settings expect for the following configurations: min CpGs per segment=5, min. delta for ASM=0.25, min confidence=0.95. Bedtools genomecov was used to overlap the bin regions identified for each individual sample to find the number of samples having ASM in each bin of the genome at the same direction of methylation differences. Only ASM bins identified in five or more samples with the same direction were kept. Post-processing filtering included requiring min. delta for ASM at single CpG resolution = 0.3 with two or more ASM CpGs within a window of 200bp (**Supplementary Figure 1**). POE-me bins were generated from the filtered CpG lists by merging imprinting ASM CpGs within 1kb.

Similarly, non-imprinted bin regions were generated by taking uninterrupted consecutive CpGs measured in at least 5 samples, separated by less than 1kb, all having ASM detected in 50% or fewer of the samples. The control set of background bins were length-matched to the detected imprinted bins by counting the imprinted regions in each length class (0-200bp/200-400bp/400-600bp/600-800bp/800-1000bp/1000+bp) and then randomly selecting, for each length class, the same number of bins from the non-imprinted bin regions of the same length class.

Putative imprinted regions were annotated regarding overlap with DNAse I hypersensitive sites^46^ (min overlap of 1bp). The webtool GREAT (version 4.0.4) was used to retrieve genes with an overlapping regulatory domain, where the regulatory domain of a gene is the region 5 kb upstream and 1 kb downstream of the transcription start site (TSS). Additionally, distal gene elements were retrieved where the regulatory domain of the genes extends up to 1 Mb.

### Gene enrichment analysis

The web-based portal Metascape^47^ (version 3.5.20250707) was used for gene list annotation and analysis using 1021 genes as the input list and default settings.

### RNA isolation and quality control

Cleaned villi from POC was stored in 1mL RNAlater Solution (Fisher Scientific, AM7020) at -70 °C. To isolate RNA, vials were slowly thawed on ice, then transferred to a biosafety cabinet and measured. Less than 30mg of tissue was used as RNA input. The tissues were suspended in 350 uL RLT buffer (Qiagen, 79216) supplemented with 10% β-mercaptoethanol (β-ME) (Sigma Aldrich, M3148-25ML) and RNA was isolated using RNeasy Mini Kit (Qiagen, 74104). The protocol provided in the kit was followed for the steps pertaining to tissues. Importantly, 30 uL of RNase free water (Qiagen, 129112) was used as the eluate. Once added, RNase free water was incubated on the column for 5 minutes at room temperature, then centrifuged at ≥ 8,000 g for 1 minute. This step was repeated with the elution, again incubated for 5 minutes, then centrifuged at ≥ 8,000 g for 1 minute to amplify the RNA concentration.

The isolated RNA was quantified with Qubit RNA Broad Range Assay kit (Invitrogen, Q10210). Then the RNA integrity number (RIN) was assessed using RNA ScreenTape (Agilent 5067-5576 and 5190-6506) on an Agilent Tapestation 2200 platform. Samples with a RIN score greater than 7 were kept as they are more likely to succeed in downstream assays.

### PacBio HiFi long-read transcript (isoform) sequencing and analysis

In total of 300ng of RNA was used as an input into cDNA synthesis and library preparation as described by PacBio in their protocol, “Preparing Kinnex libraries using the Kinnex full-length RNA kit,”. Iso-seq primer barcodes were used during cDNA amplification to allow for multiplexing in sequencing. To target longer fragments, SMRTbell cleanup beads (PacBio, 102-158-300) were added (0.86x), as specified in the protocol. cDNA concentration was evaluated using Qubit dsDNA High Sensitivity Assay kit (Fisher Scientific, Q32856) and equal amount of cDNA was pooled for a total of 55 ng for Kinnex PCR. Library preparation was completed following the protocol mentioned above. Final quality control was analyzed by diluting finished Kinnex library, 1:10, and recording concentration using Qubit dsDNA High Sensitivity Assay kit (Fisher Scientific, Q32856). To determine the final size prior to sequencing, the library was diluted to 250 pg and analyzed on Agilent Femto pulse.

The IsoSeq3 pipeline (https://github.com/PacificBiosciences/IsoSeq) was used to generate a non -redundant set of unique isoforms. In short, full-length non-concatemer (FLNC) were clustered into isoforms, and aligned to the reference genome (GRCh38) using pbmm2 with the argument --preset ISOSEQ. The aligned transcripts collapsed into a non-redundant set of isoforms based on exonic structures as described in the Gencode v39 annotation. Finally, the isoforms were classified and filtered using Pigeon using default parameters. Isoform-specific allele counts were determined for genes of interest, based on results from both Kinnex long-read RNA sequencing. A custom R script (R version 4.4.0) was written to sort reads with transcription start sites in distinct user-specified ranges into separate bam file outputs. Secondary splits were also made based on the presence or absence of other exons, when such splicing variation occurred in a potentially informative number of reads. SNPs and SVs present in the transcripts were associated with WGS from probands and parents when available, allowing both maternal and paternal allele counts to be made for each isoform or isoform group. Parental genomic data also made possible the identification of maternal tissue contamination in sample types (i.e. placental) where that was a possibility. When parental data was not present and variants could not be assigned as maternal or paternal, alleles were defined as an arbitrary parent 1 and parent 2. Genes with no informative variants were measured for overall coverage per isoform. Overall coverage, SNP and SV identification, variant read counts, and visualization were performed for the separated bam files using the Integrative Genomics Viewer (IGV, version 2.17.4).

### Integration of GWAS summary statistics

FinnGen GWAS summary statistics (release R12) were obtained from the relevant FinnGen Google Cloud storage bucket (see **Data Availability**). Each GWAS file was intersected with methylation and background bins (plus a window +/- 10Kb around bins) using bedtools^48^ (version 2.30.0) (window flag, -w 10000 parameter). GWAS phenotypes were groups into categories using data from the metadata file (“finngen_R12_manifest.tsv”), retaining categories with >=25 distinct GWAS phenotypes. Enrichment for phenotype categories with variants mapping to methylation compared to background bins was performed using Fisher’s exact test for categories with >=50 variants with GWAS P-value <= 1e-08 and beta>0 (i.e. risk variants) mapping to either methylation or background bins. GWAS peaks and overlapping methylation bins were visualized using LocusZoom (available at https://my.locuszoom.org/). deCODE/UK Biobank GWAS meta-analysis summary statistics for birth weight and birth length were obtained from the deCODE server (see **Data Availability**). Each GWAS file was intersected with methylation and background bin sets using bedtools^48^ (version 2.30.0) (intersect flag). Differences in the proportion of overlap between methylation and background bins with GWAS summary statistics were assessed using Fisher’s exact test. We further assessed differences in birth weight for maternally or paternally inherited variants using phenotypic data from GA4K and DDD (see **Phenotype data**). Variants overlapping birth weight GWAS variants were extracted from the imputed genotype callsets for each respective study using plink^49^ (version 2) (--extract flag), further subsetting to full trios (proband, mother, father). Variants were tested for differences in birth weight by maternal/paternal inheritance in a linear regression model adjusting for proband sex (both GA4K and DDD) and age at measurement (GA4K only).

### Variant Classification in DDD

Variants were extracted from the Deciphering Developmental Disorders (DDD) study dataset EGAD00001004389 (Total N=28475 samples, 9859 probands with parents) and initially filtered for PASS variants, for the provide Maximum Allele Frequency across 1000G, UK10K, ESP and DDD MAFs (calculated by DDD as MAX_AF) < 0.001 or not having a MAX_AF, and then filtered using the provided annotations (from ensembl VEP) for missense and LOF consequences: ‘transcript_ablation’, ‘splice_acceptor_variant’, ‘splice_donor_variant’, ‘stop_gained’, ‘frameshift_variant’, ‘stop_lost’, ‘start_lost’, ‘transcript_amplification’, ‘feature_elongation’, ‘feature_truncation’, ‘missense_variant’.

A list (“pathogenic patient variant list”) was generated of the above variants reported in probands (and parents) (n=10481 individuals comprising 3569 probands plus parents) reported by DECIPHER by 4 April 2022 annotated with clinical and automated pathogenicity assertions^31^. Similarly, another list (“non-pathogenic patient variant list”) was generated of the above variants observed in all the other probands and their parents (n=18118 comprising 6290 probands and where 268 parents are shared with the pathogenic patients’ parents). Variants were further filtered for GnomAD v4.1 MAF < 1×10^−5^ and annotated with hg19 AlphaMissense^31^ (2025-05-15). For each variant, we reported whether a variant (genotype 0/1) was also observed (genotype 0/1) in only the mother or the father, with the other parent as ref (genotype) 0/0 (putatively transmitted from that parent) or whether the variant appeared to be *de novo* (both parents genotype 0/0). Taking the 5852 POE-me bins, crossmap was used to translate the coordinates to hg19 for a total of 5830 successfully remapped bins. In total 4807 of these regions were within 20kb of one or more genes from the UCSC knownCanonical (Gencode V45, Oct 2024) (4317 maternal regions and 490 paternal regions). Each knownCanonical gene definition was annotated with the number of overlapping maternal regions and the number of overlapping paternal regions and then intersected with the pathogenic and non-pathogenic variants lists. Genes assessed in the equilibrium analysis were required to have exclusively maternal, or exclusively paternal, overlapping bins and with >=2 variants for each analysis of pathogenic or benign.

### Variant lookup in GA4K and CGGL Databases

Variants in 34 candidate genes identified in DDD analysis were extracted from 10456 GA4K and 24027 CGGL exomes or genomes from probands and parents stored in Xetabase filtered for GnomAD Genomes v3.1.2 alt frequency < 1×10^−5^ and manually annotated with hg38 AlphaMissense (2025-05-15). For each variant, we reported whether a variant (genotype 0/1) was also observed (genotype 0/1) in only the mother or the father, with the other parent as ref (genotype) 0/0 (putatively transmitted from that parent) or whether the variant appeared to be *de novo* (both parents genotype 0/0).

### Rare disease gene annotation

Rare disease gene annotations were obtained from OMIM and DDG2P (**Data Availability**). OMIM genes were categorized into autosomal dominant (AD), autosomal recessive (AR), or both (AD/AR) based on free text match and negation using the “phenotypes” column. DDG2P gene annotations were filtered to those with “strong” or “definitive” labels (“confidence_category” column), further retaining genes with “biallelic_autosomal” or “monoallelic_autosomal” annotations only (“allelic_requirement” column).

### Phenotype data

Height and weight observation data for GA4K, including measure date/time, were obtained from the electronic medical record at the GA4K study site at Children’s Mercy Hospital. R package cdcanthro (version 0.1.1) was used to calculate age and sex adjusted Z-scores for height and weight using CDC guidelines. Further, BMI Z-scores were calculated using the formula Weight/(Height/100)2. The average (median) Z-score for each measurement was computed for samples with more than one observation time point. DDD birth weight Z-scores was sourced from dataset EGAD00001010136 (see **Data Availability**) (specifically, “birthweight_sd” column in file “DDD_growth_data_2025-03-24.csv”).

### Genotype imputation and filtering

Genotyping array data for two rare disease cohorts (GA4K and DDD) were used in analyses of variant effects stratified by maternal/paternal inheritance. Genotyping of the full available GA4K study cohort was performed using the Avera Global Screening Array (24v1-0_A1, stranded) (**Data Availability**). Genotyping variant calls in hg38 build for samples with genotyping call rate >= 90% were then used as input for imputation using the TOPMed imputation server with the r3 (1.0.0) reference panel^50^. Variants were filtered to those with imputation quality R^2^ ≥L0.8. The final proband cohort comprised 1434 female and 1625 male samples. Genotyping data from the DDD study (Data Availability), comprising 571 female and 893 male samples (dataset EGAD00010002568; hg38 genome build) were used as input for imputation using the TOPMed imputation server with r3 (1.0.0) reference panel as above. Variants were filtered to those with imputation quality R^2^L≥L0.8 and which were also available in the filtered GA4K imputed callset (N variants = 23,952,791).

### Hypermethylation Outlier Analysis

Discovery of hypermethylation outliers among rare disease probands and family members (n=1,112) was based on tiled z-score similar to what we earlier described in smaller dataset^13^. Briefly, z-score distribution based on the published subset of 112 high coverage samples was used to find CpGs (min 10X read depth) where “hyper CpGs” were called when at least 3 SD outside the q5 to q95 range, this process is done for total and haploid methylation for each sample. “Hyperbins” were 200bp regions harboring a minimum of 2 hyper CpGs in same bin in same haplotype or total methylation. We then averaged all z-scores within hyper bins and kept bins with average z-scores > 3 after averaging. We required that there were no more than 4 families harboring the “hyper bin” among the sample (less than <1% frequency). Among the probands a total 64,321 rare hyper bins (∼100 per proband) were observed. For “de novo” methylation outlier discovery we focused on the of hyper bins that were fully informative for 5mC-HiFi-GS methylation in both parents (n=5,636) to describe the “de novo” hypermethylation requiring deviation of z>3 from both parents. In total we found 1,920 de novo methylation bins of which 171 also overlapped OMIM 5’ proximal region (+/- 10kb).

## Ethics Declaration

The prenatal and GA4K studies fall under a protocol approved by the Children’s Mercy Institutional Review Board (Study No. 11120514). Informed written consent was obtained from all participants before study inclusion. Participants were not compensated for study participation. Salvaged semen samples were deidentified and considered non-human subjects research as determined by the Children’s Mercy Institutional Review Board (Study No. 16020084). Studies of patients who have had clinical genetic testing through Children’s Mercy for reporting diagnostic yield, genotype phenotype correlations, novel variants, types of diagnoses, and technical metrics of genetic testing was approved by the Children’s Mercy Institutional Review Board (Study No. 00000175).

## Data availability

The 5-base HiFi-GS, and HiFi long-read transcript sequencing (IsoSeq) generated in this study have been deposited in the dbGAP (https://www.ncbi.nlm.nih.gov/gap/) database under accession code phs002206.v5.p1 [https://www.ncbi.nlm.nih.gov/projects/gap/cgi-bin/study.cgi?study_id=phs002206.v5.p1]. Raw and processed data are available under restricted access due to IRB regulations and informed consent limiting access to users studying genetic diseases. Data access is provided by dbGAP (https://dbgap.ncbi.nlm.nih.gov/aa/wga.cgi?page=login) for certified investigators with local IRB approval in place. The reference genome GRCh38 (GCA_000001405.15) used in this study is available at ftp://ftp.ncbi.nlm.nih.gov/genomes/all/GCA/000/001/405/GCA_000001405.15_GRCh38/seqs_for_alignment_pipelines.ucsc_ids/GCA_00001405.15_GRCh38_no_alt_analysis_set.fna.gz. WGS and HTG sequences that are not part of the human reference genome, GRCh38 (including the ALT sequences) was added using https://www.ncbi.nlm.nih.gov/assembly/GCA_000786075.2/. Gene annotation data is available from GENCODE (https://www.gencodegenes.org/). The GnomAD v4.1.0 joint allele frequency vcf data used to annotate the DDD dataset is available at https://gnomad.broadinstitute.org/data. Xetabase v3.0.0 GnomAD Genomes v3.1.2 annotations were taken from CellBase 5.8.2 . DDG2P disease gene annotations are available at https://www.ebi.ac.uk/gene2phenotype/downloads/DDG2P.csv.gz. OMIM disease gene annotations are available at https://www.omim.org/downloads/. deCODE/UKBB GWAS meta-analysis summary statistics are available at https://download.decode.is/form/2021/Birthweight2021.gz (birth weight) and https://download.decode.is/form/2021/Birthlength2021.gz (birth length). Deciphering Developmental Disorders (DDD) data is available from the European Genome-Phenome Archive at the European Bioinformatics Institute using study accession number EGAS00001000775. FinnGen R12 GWAS summary statistics are available at https://www.finngen.fi/en/access_results.

## Code availability

Only publicly available tools were used in data analysis as described wherever relevant in Methods.

## Acknowledgments

We would like to thank all patients and families for participating in the Genomic Answers for Kids study. We also want to acknowledge the participants and investigators of the FinnGen study. This work was made possible by the generous gifts by the Sosland Foundation and the Edgerly Family Foundation as well as gifts to the Genomic Answers for Kids program at Children’s Mercy Kansas City. We also would like to thank Rebecca McLennan, Nick Nolte, Dan Louiselle and Alyse Peters for their work in project management, sample processing and handling; Margaret Gibson, Shanna Beyer, Laura Puckett and Adam Walters for their work in library preparation and sequencing and the clinical coordination team including Bradley Belden, Amy Ross, Emily Munden, Grace Baker and Victoria Sierant for their work in clinical coordination. The DDD study presents independent research commissioned by the Health Innovation Challenge Fund (grant number HICF-1009-003), a parallel funding partnership between Wellcome and the Department of Health, and the Wellcome Sanger Institute (grant number WT098051). The views expressed in this publication are those of the authors and not necessarily those of Wellcome or the Department of Health. The study has UK Research Ethics Committee approval (10/H0305/83, granted by the Cambridge South REC, and GEN/284/12 granted by the Republic of Ireland REC). The research team acknowledges the support of the National Institute for Health Research, through the Comprehensive Clinical Research Network. T.P holds the Dee Lyons/Missouri Endowed Chair in Pediatric Genomic Medicine and E.G holds the Roberta D. Harding & William F. Bradley, Jr. Endowed Chair in Genomic Research. The research was supported by the National Institutes of Health: HD115359 (E.G., T.P.), HD117565 (E.G., T.P.), GM146966 (C.S.), HG012422 (C.S.).

## Author Contributions

E.G., and T.P. conceived and designed the study; I.T., E.R., J.H., M.L., K.L., C.M., prepared, provided and/or analyzed clinical samples and associated data; W.A.C., B.K., A.F.J., C.B., C.S-S., provided bioinformatics support; C.S., E.G., and T.P. analyzed the data and interpreted results of experiments; C.S., E.G., and T.P. prepared figures and drafted manuscript; All authors approved the final version of manuscript.

