## Supplemental Figures for "Expanded map of genomic imprinting reveals insight into human disease"

### Table of contents


| | |
|---|---|
| Supplementary Figure 1 | Page 2 |
| Supplementary Figure 2 | Page 3 |
| Supplementary Figure 3 | Page 4 |
| Supplementary Figure 4 | Page 5 |
| Supplementary Figure 5 | Page 6 |
| Supplementary Figure 6 | Page 7 |
| Supplementary Figure 7 | Page 8 |
| Supplementary Figure 8 | Page 9 |
| Supplementary Figure 9 | Page 10 |
| Supplementary Figure 10 | Page 11 |
| Supplementary Figure 11 | Page 12 |
| Supplementary Figure 12 | Page 13 |
| Supplementary Figure 13 | Page 14 |
| Supplementary Figure 14 | Page 15 |
| Supplementary Figure 15 | Page 16 |
| Supplementary Figure 16 | Page 17 |
| Supplementary Tables 1-15 | .xlsx |

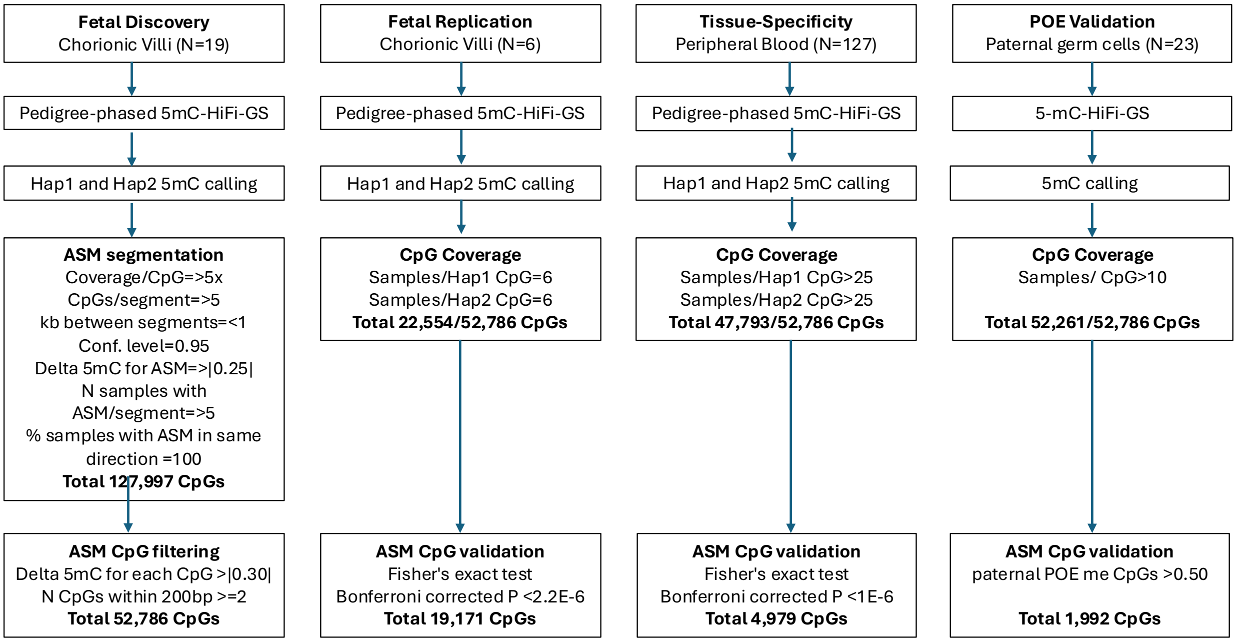


**Supplementary Figure 1.** Flowcharts outlining data analysis steps for discovery, replication, tissue-specificity, and POE validation arms of the study.

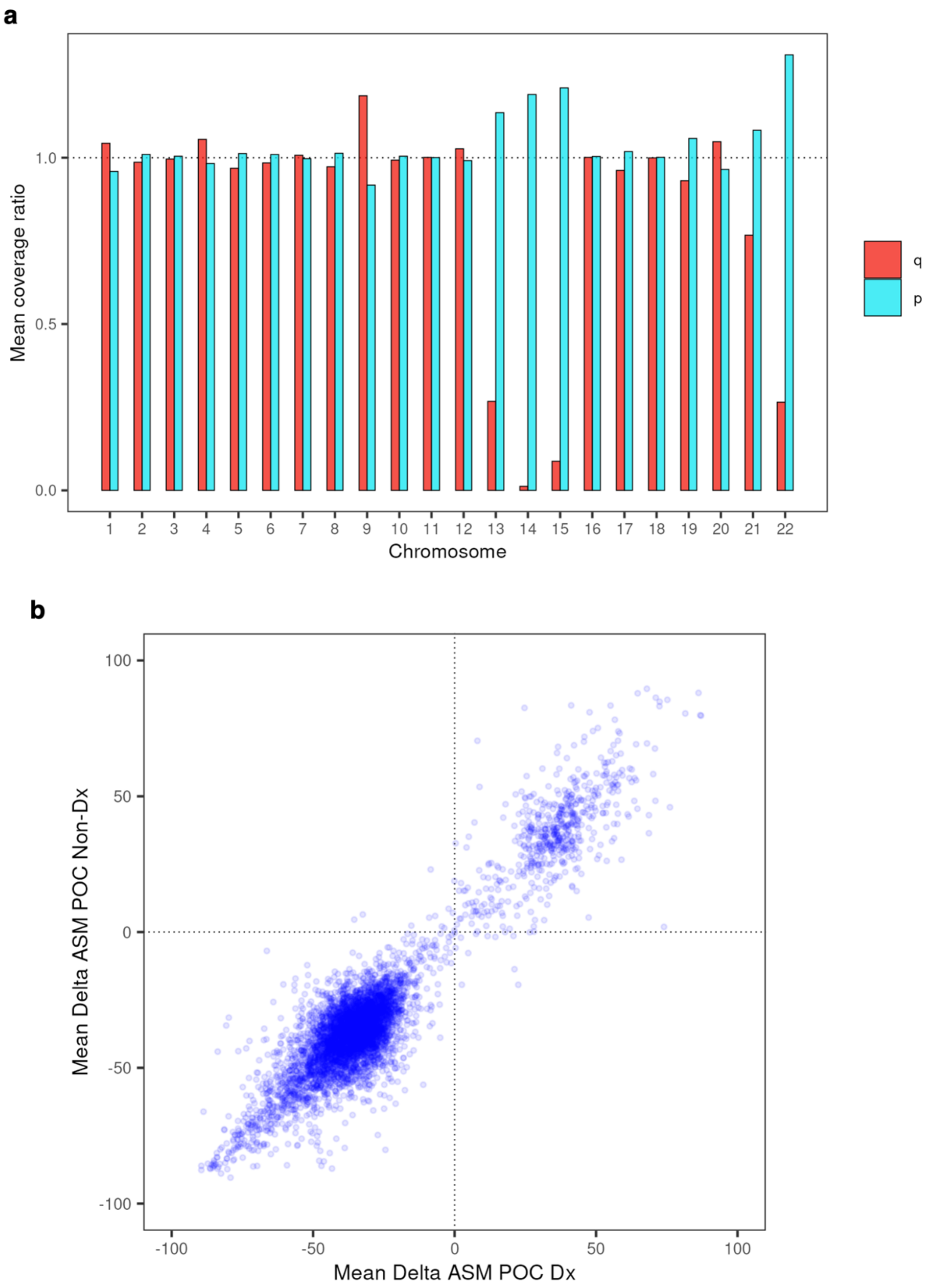


**Supplementary Figure 2. a.** Mean coverage ratio for each POE-me bin across p and q arms of chromosomes. **b.** Correlation of ASM difference in the POE-me bins in fetal samples diagnosed with aneuploidy vs. chromosomal normal samples.

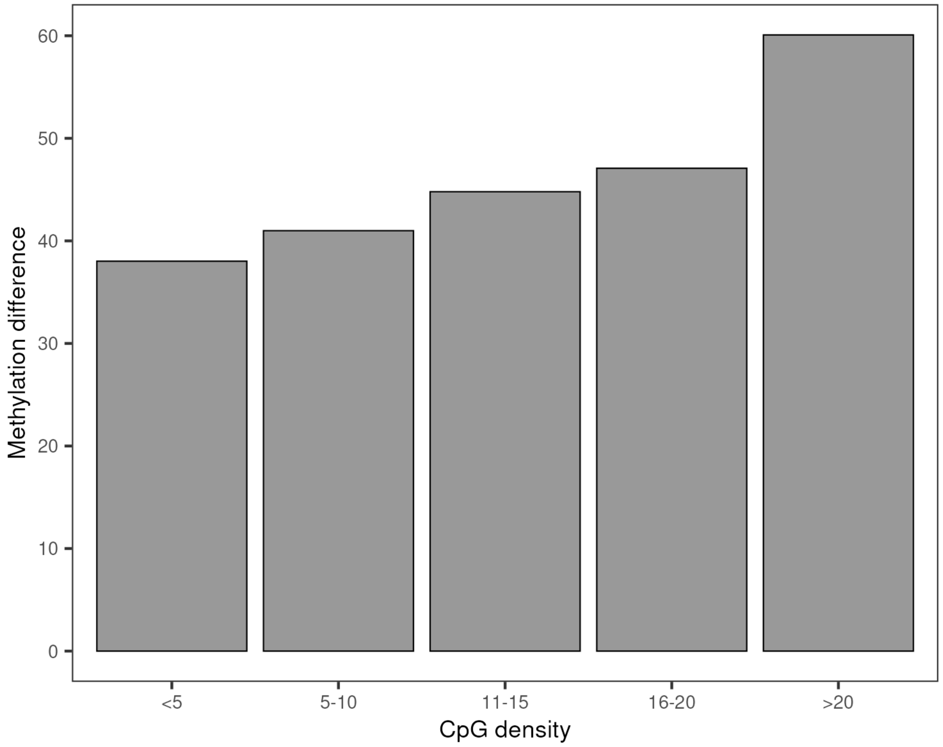


**Supplementary Figure 3.** Methylation difference as a function CpG density.

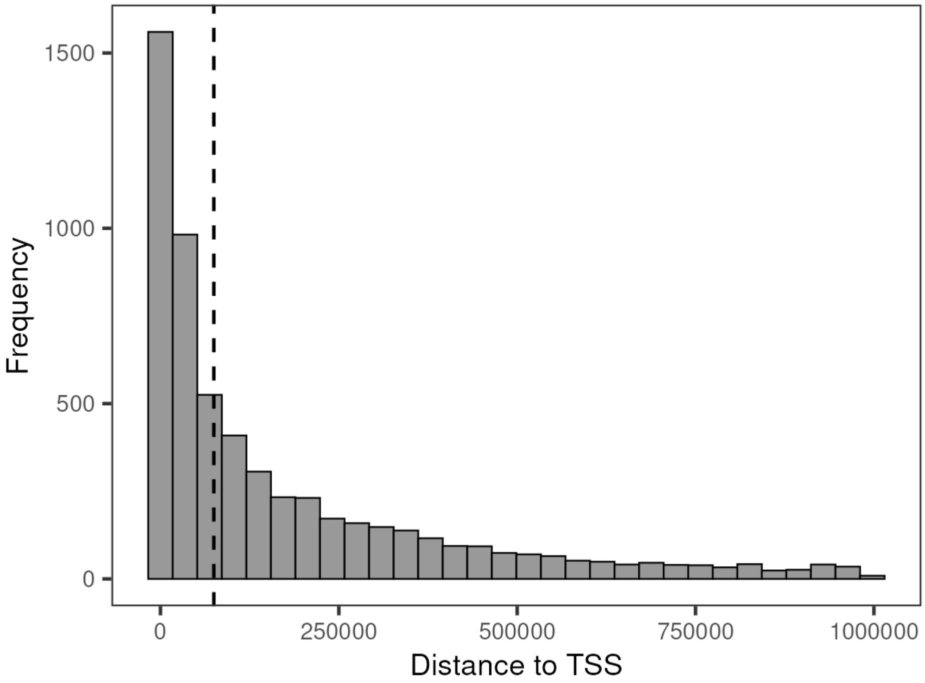


**Supplementary Figure 4.** Histogram of distance to transcription start site (TSS) for each POE-me bin as determined by GREAT.

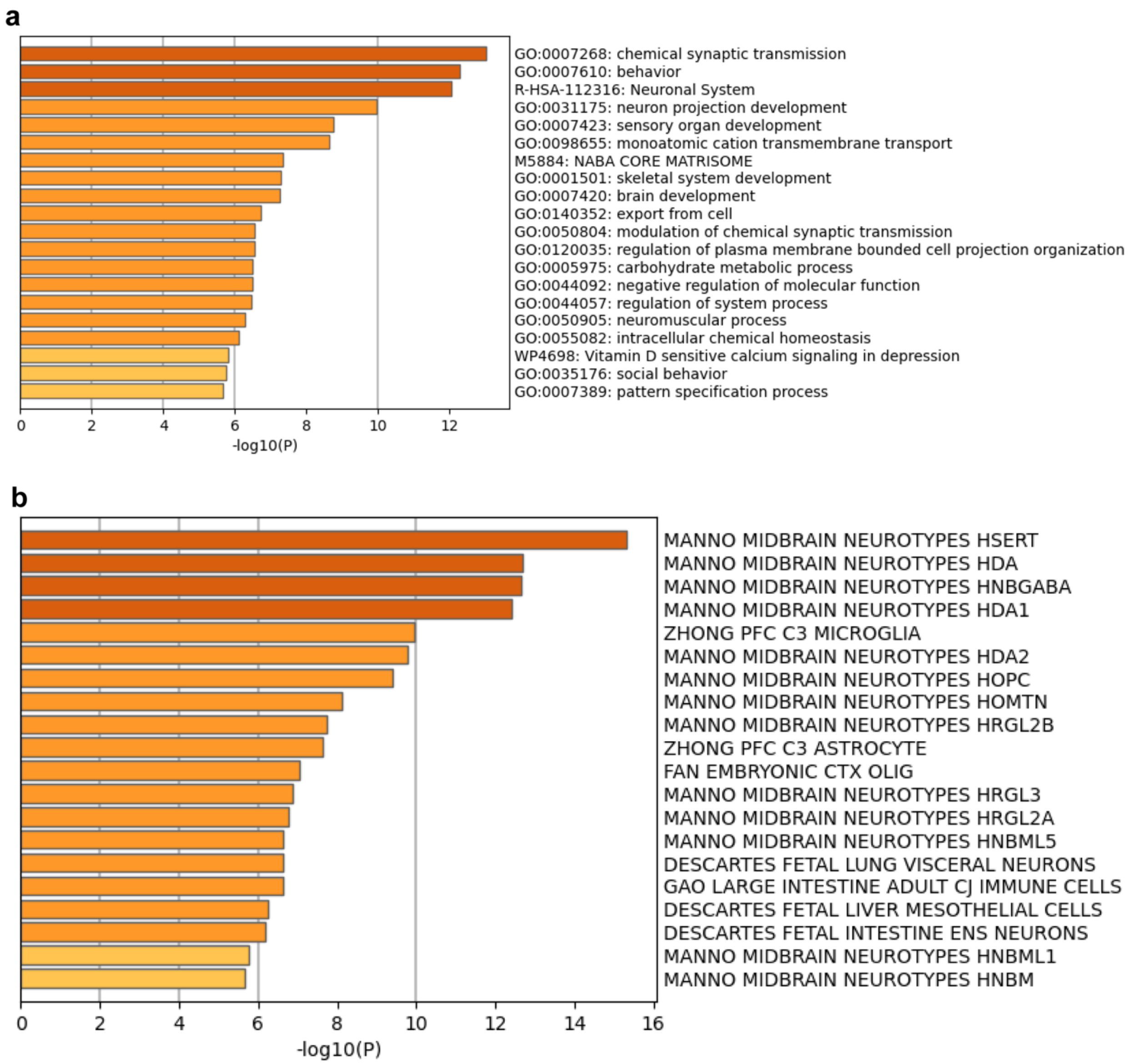


**Supplementary Figure 5. a.** Bar graph of enriched ontology clusters for genes mapping to POE-me bins based on statistical significance (-log10 p-value, x-axis). **b.** Bar graph of enriched cell types for genes mapping to POE-me bins based on statistical significance (-log10 p-value, x-axis).

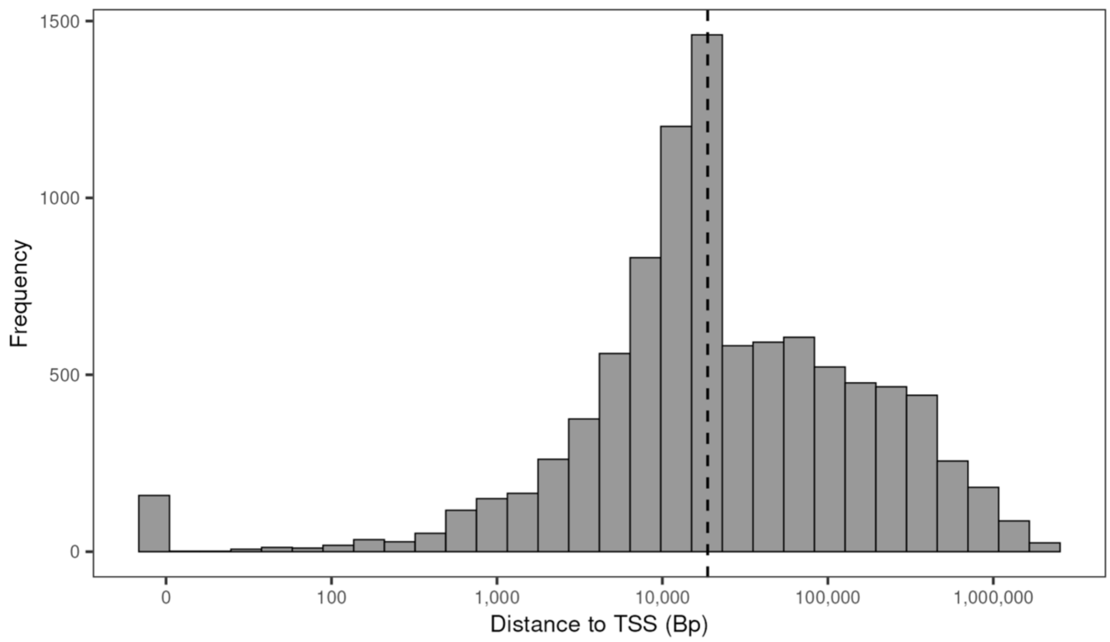


**Supplementary Figure 6.** Histogram of distance to transcription start site (TSS) for each POE-me bin to any GENCODE V45 protein coding or long non-coding transcript**.**

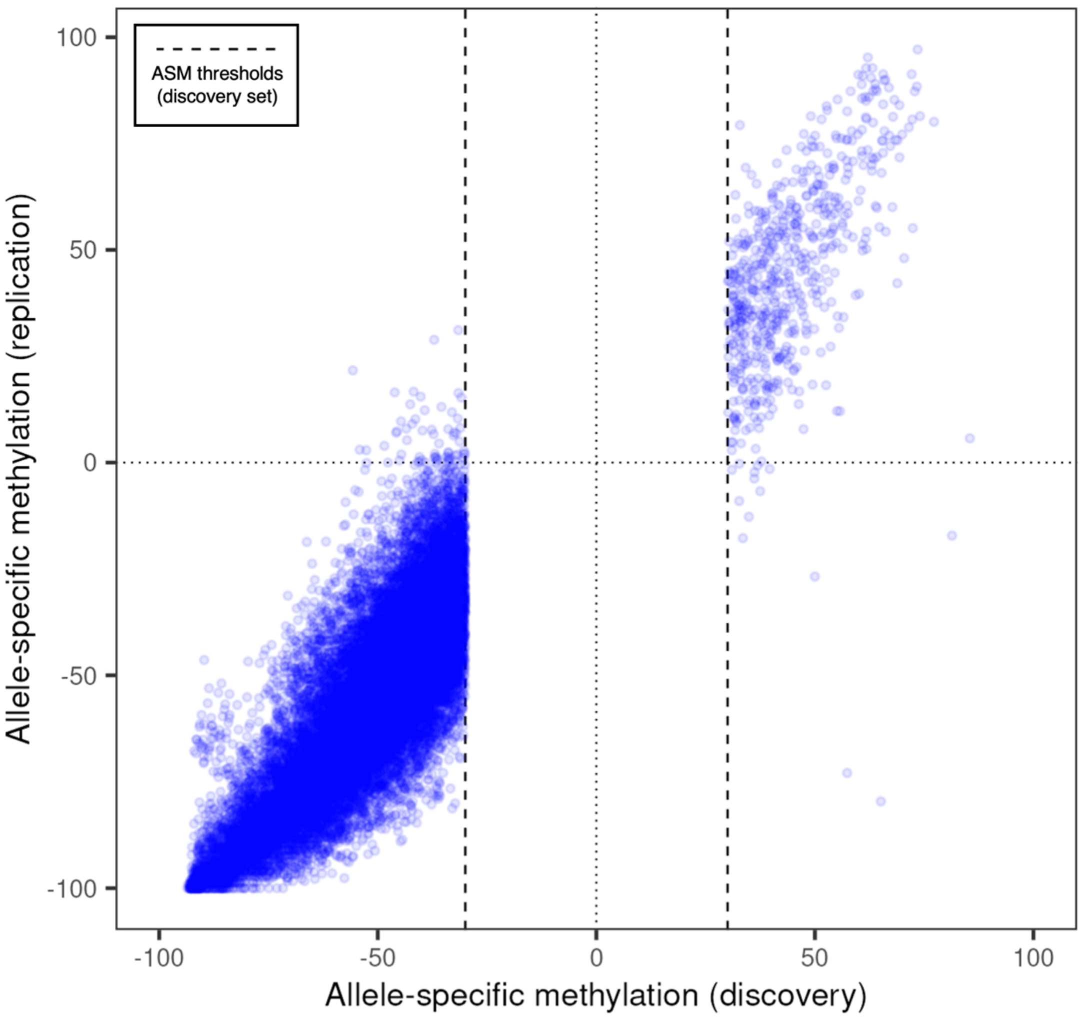


**Supplementary Figure 7.** Scatterplot of allele-specific methylation (ASM) in discovery and replication cohorts. Vertical dashed lines indicate ASM thresholds used in discovery cohort.

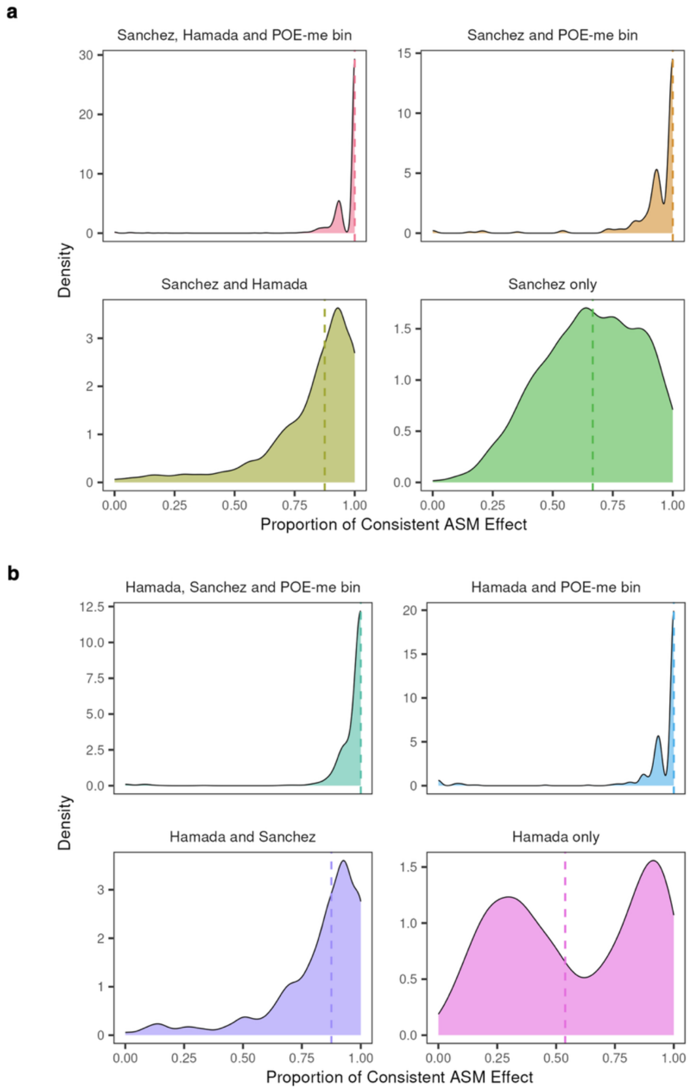


**Supplementary Figure 8. a.** Distribution of the proportion (x-axis) of fetal samples showing same direction of ASM as determined by haplotype-resolved methylation profiles of suggested imprinting regions presented by Sanchez-Delgado et al[18] for regions validated by: Hamada et al[17] and POE-me (upper left), POE-me bins only (upper right), Hamada et al[17] only (lower left) or no additional study (lower right). **b.** Distribution of the proportion (x-axis) of fetal samples showing same direction of ASM as determined by haplotype-resolved methylation profiles of suggested imprinting regions presented by Hamada et al[17] for regions validated by: Sanchez-Delgado et al[18] and POE-me (upper left), POE-me bins only (upper right), Sanchez-Delgado et al[18] only (lower left) or no additional study (lower right).

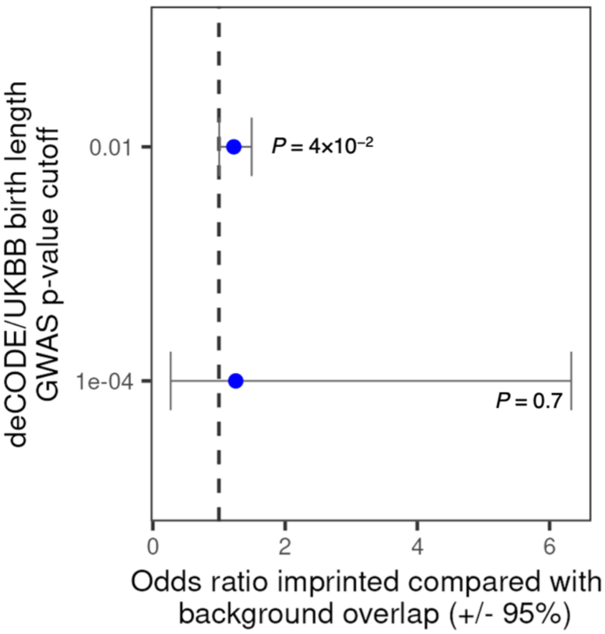


**Supplementary Figure 9.** Odds ratios comparing methylated and background bins overlapping birth length GWAS hits, stratified by GWAS P-value.

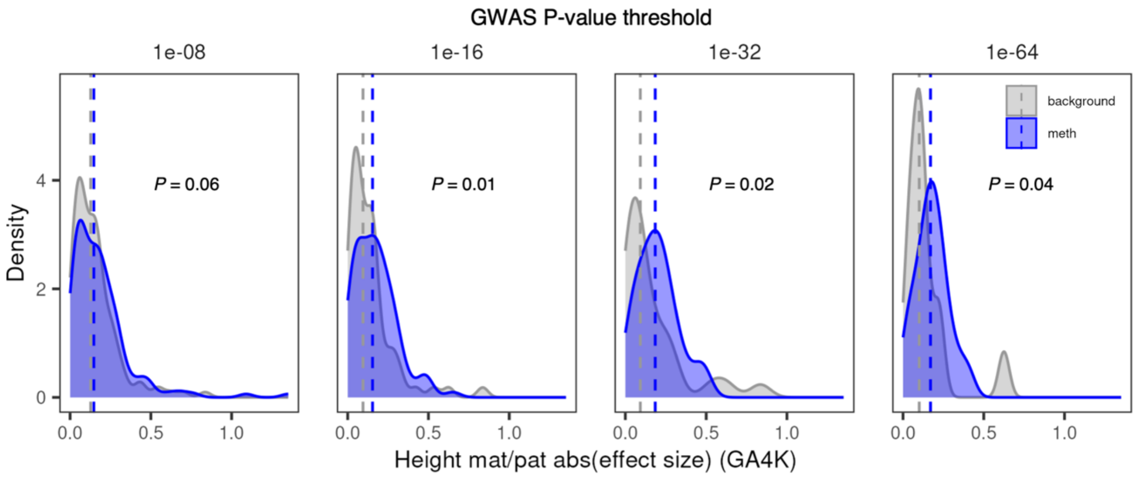


**Supplementary Figure 10.** Logistic regression coefficients comparing effect on height in GA4K probands of paternally or maternally inherited variants for methylated (blue) and background (gray) bins, stratified by birthweight GWAS P-value.

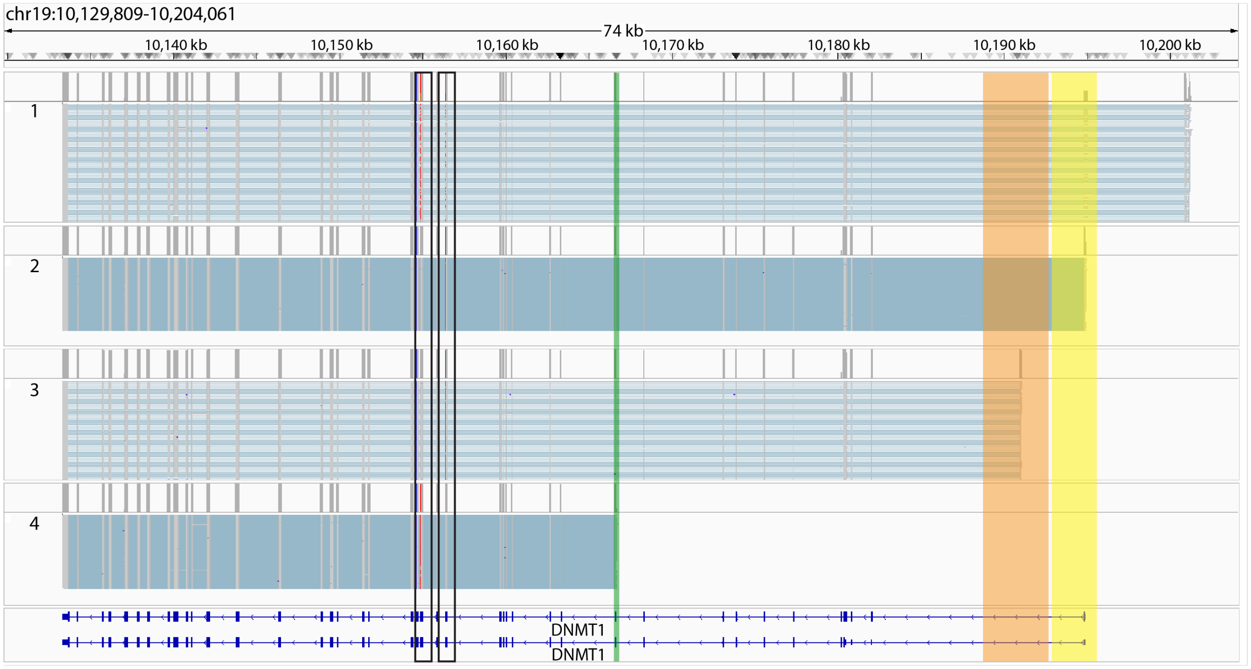


**Supplementary Figure 11.** Long-read isoform profiling in fetal chorionic villi samples of the *DNMT1* locus (chr19:10,129,809-10,204,061) detecting four major isoforms: 1) biallelic expressed oocyte specific variant (DNMT1o isoform), 2) paternally expressed somatic isoform (DNMT1s isoform) linked to known maternally imprinted region (yellow box), 3) paternally expressed isoform linked to novel maternal POE-me bin (orange box) and 4) maternally expressed isoform linked to novel paternal POE-me bin (green box). Black boxes indicate two informative genetic variants used for ASE analysis.

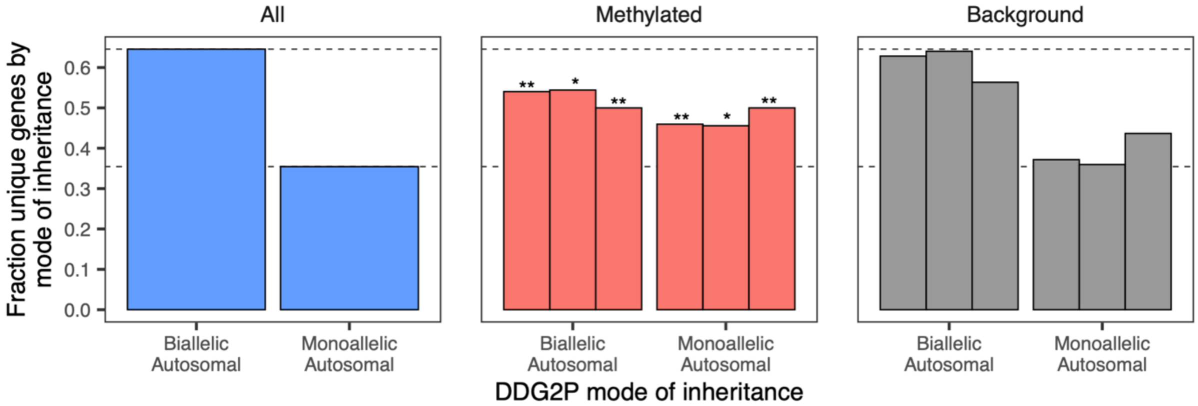


**Supplementary Figure 12.** Fraction of genes for each indicated mode of inheritance in DDG2P (blue), DDG2P genes overlapping methylation bins (red), and DDG2P genes overlapping background bins (blue). Methylation and background bins are stratified by bin size (from left to right: any bin; >=100 bp; >=250 bp).

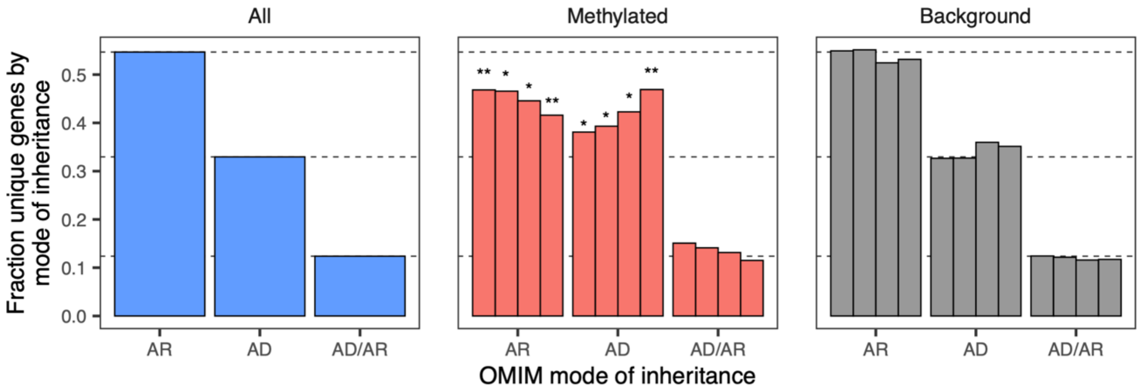


**Supplementary Figure 13.** Fraction of genes for each indicated mode of inheritance in OMIM (blue), OMIM genes overlapping methylation bins (red), and OMIM genes overlapping background bins (blue) after removing known imprinted genes. Methylation and background bins are stratified by bin size (from left to right: any bin; >=100 bp; >=250 bp; >=500 bp).

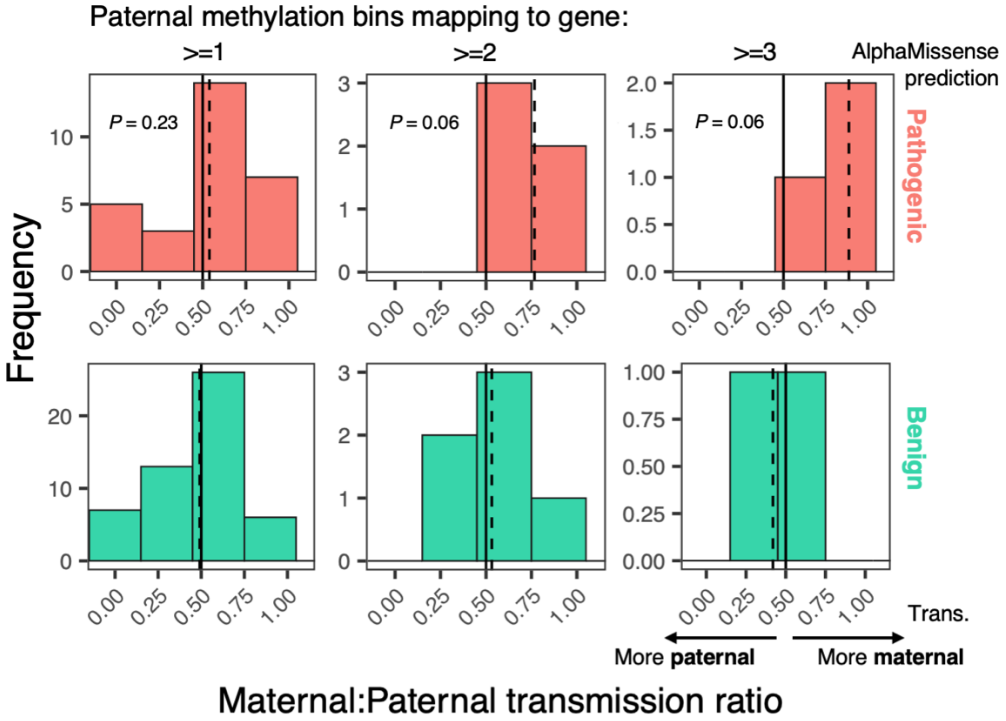


**Supplementary Figure 14.** Maternal-to-paternal transmission ratios for predicated pathogenic (top; red) and benign (bottom; green) variants in genes overlapping paternal methylated bins. Results are stratified by number of bins mapping to a gene (from left to right: >=1 bin; >=2 bins; >=3 bins).

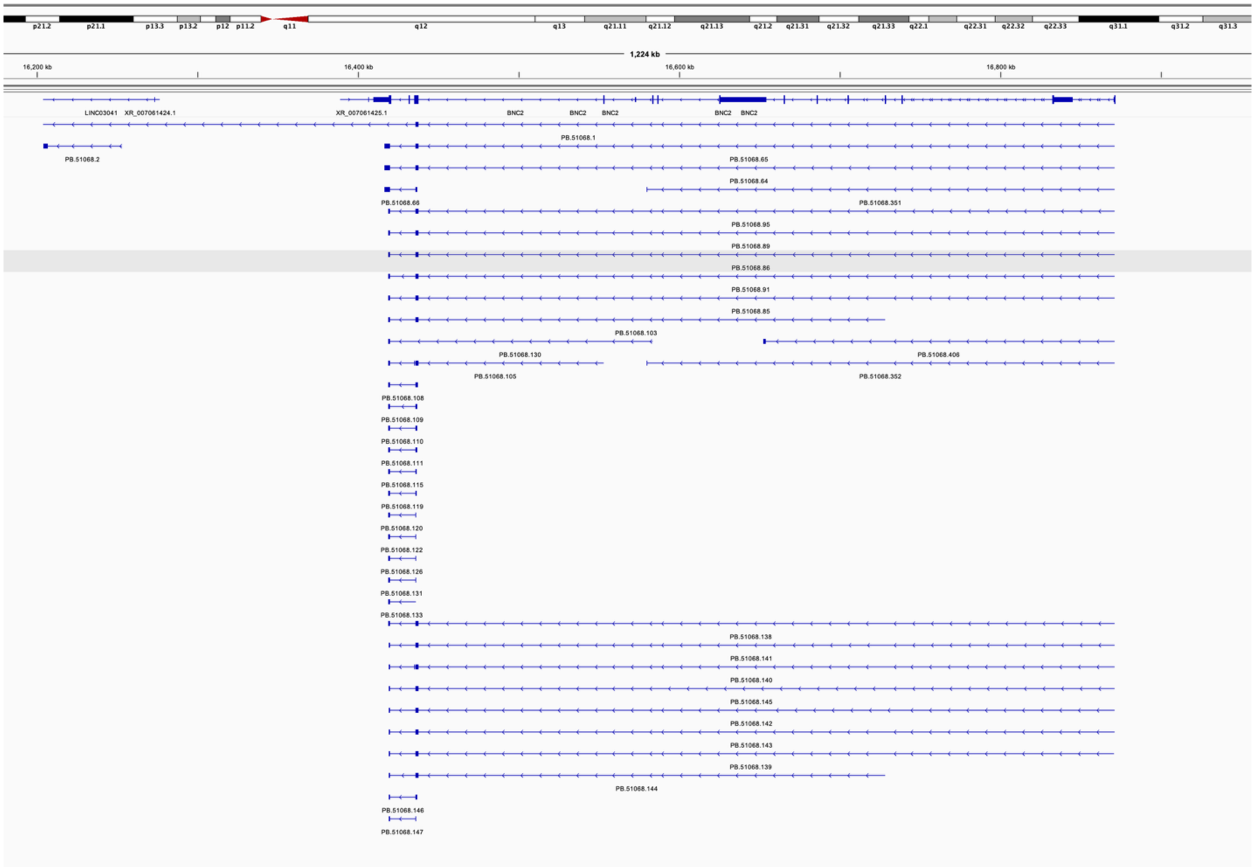


**Supplementary Figure 15.** IGV visualization of *BNC2* isoforms identified in fetal chorionic villi samples by long-read RNA sequencing. Black box denotes POE-me bin.

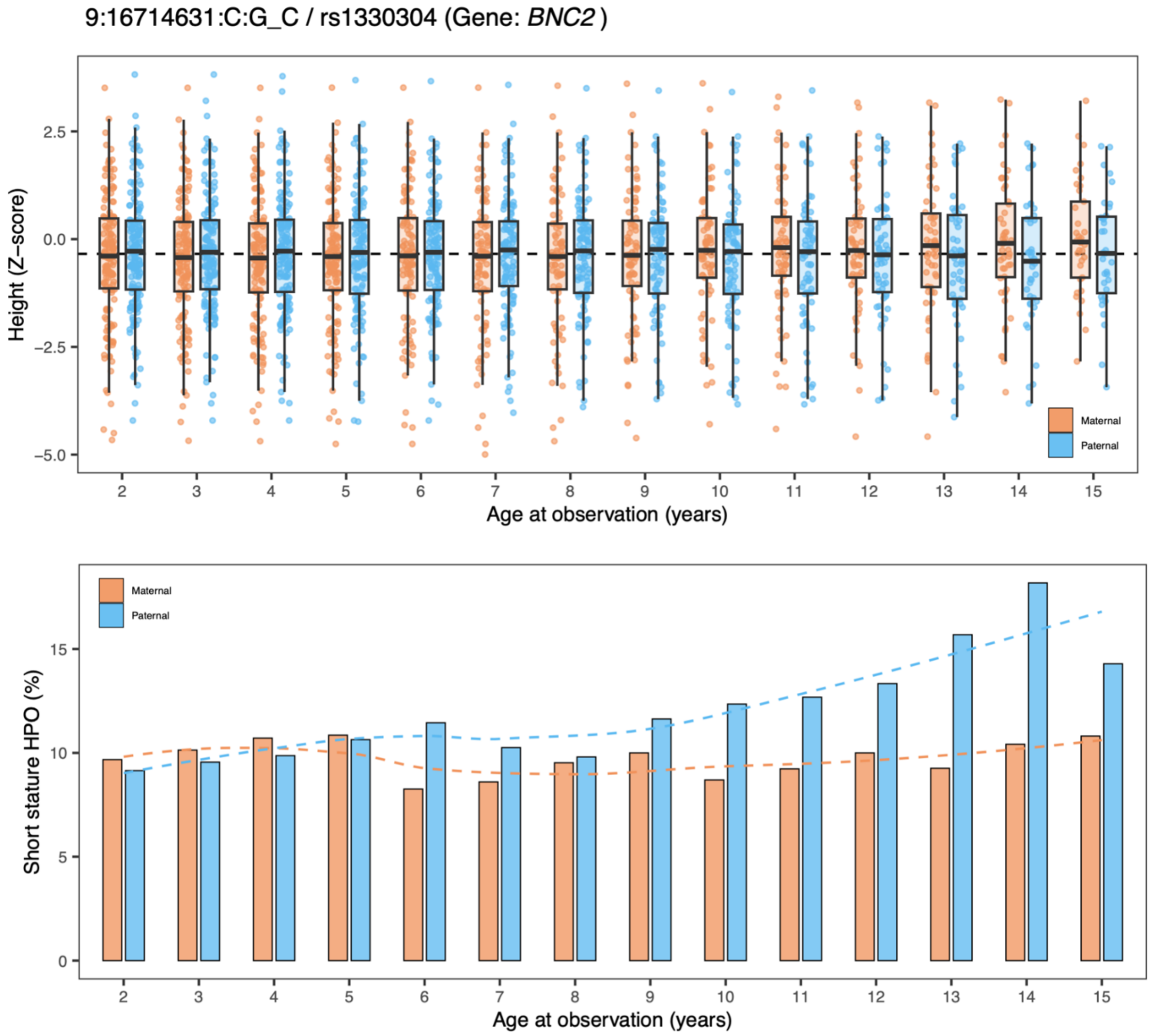


**Supplementary Figure 16.** Trajectories of height (top) and incidence of short stature (bottom) across age in years at observation for maternal or paternal inheritance of a previously associated variant for height.
